# Global estimates of the fitness advantage of SARS-CoV-2 variant Omicron

**DOI:** 10.1101/2022.06.15.22276436

**Authors:** Christiaan van Dorp, Emma Goldberg, Ruian Ke, Nick Hengartner, Ethan Romero-Severson

## Abstract

New variants of SARS-CoV-2 show remarkable heterogeneity in their relative fitness both over time and space. In this paper we extend a previously published model for estimating the selection strength for new SARS-CoV-2 variants to a hierarchical, mixed-effects, renewal equation model. This formulation allows us to globally estimate selection effects at different spatial levels while controlling for complex patterns of transmission and jointly inferring the effects of unit-level covariates in the spatial heterogeneity of SARS-CoV-2 selection effects. Applying this model to the spread of Omicron in 40 counties finding evidence for very strong (64%) but very heterogeneous selection effects at the country level. We further considered different measures of vaccination levels and measures of recent population-level infection as possible explanations. However, none of those variables were found to explain a significant proportion of the heterogeneity in country-level selection effects. We did find a significant positive correlation between the selection advantage of Delta and Omicron at the country level, suggesting that region-specific explanatory variables of fitness differences do exist. Our method is implemented in the Stan programming language, can be run on standard commercial-grade computing resources, and should be straightforward to apply to future variants.

## Introduction

Since its emergence, the SARS-CoV-2 virus has continuously generated genetic variants that increase its transmission, e.g., D614G and variants of concerns (VOCs) such as Alpha and Delta, causing waves of COVID-19 surges across the globe (Korber et al. 2020; Davies et al. 2021; Volz et al. 2021; Dhar et al. 2021). Omicron, first detected in November 2021 in South Africa and Botswana, quickly spread globally, displaced Delta, and became the dominant variant in most countries. Initial studies on data from South Africa suggested that Omicron has a substantially higher transmission fitness than Delta, especially in individuals recovered from COVID-19 (Pulliam et al. 2021; Viana et al. 2021; Pearson et al. 2021). Experiments measuring plasma neutralization activity from vaccinated individuals demonstrated lower neutralizing antibody activities against Omicron even in fully vaccinated individuals (Schmidt et al. 2022; Collie et al. 2022; Cele et al. 2021; Lu et al. 2021; Planas et al. 2021); however, a booster shot increased neutralizing antibody activity substantially (Schmidt et al. 2022; Nemet et al. 2022; Planas et al. 2021). This suggests that Omicron is able to evade immunity induced in vaccinated individuals, providing a likely explanation for the rapid surge in Omicron in countries where a large proportion of individuals are fully vaccinated. Previous work has also quantified the reproductive number or relative growth advantage of Omicron in particular countries or settings, showing it to exceed that of previous emerging variants (Pearson et al. 2021; Weil et al. 2022) and to be greater among vaccinated individuals (Paton et al. 2022).

Although it is clear Omicron has higher transmission fitness than Delta, we still lack a global perspective on the magnitude of this advantage and its heterogeneity among countries. Analyses of other SARS-CoV-2 variants have shown large differences in their relative fitness among different countries and US states (van Dorp et al. 2021; Figgins and Bedford 2021). It is expected the transmission advantage of Omicron is also heterogeneous across countries, due to the difference in the circulating variants in each country, the different immunological landscapes arising from natural infection and vaccination, the different non-pharmaceutical intervention strategies adopted in different countries, and potentially many other factors. Unknown, however, is the extent to which among-country heterogeneity is actually explained by any known factors.

To quantify the transmission advantage of Omicron across many countries and explore possible causes of its heterogeneity, we analyzed case count and variant frequency data with a renewal model (as Figgins and Bedford 2021) that is hierarchical across countries (as Paton et al. 2022; van Dorp et al. 2021) and includes country-level covariates (as Paton et al. 2022), while allowing for a shorter generation time of Omicron (which could otherwise bias the results, cf. Pearson et al. 2021). We found that across forty countries, the substantial heterogeneity in Omicron’s fitness could not be explained by immunological-related covariates. It did, however, show some correlation with fitnesses for Delta, suggesting that fixed country-level effects may exist. We additionally demonstrate the efficacy of a hierarchical model structure in quantifying overall variant fitness.

## Methods

### Data

Variant counts per country per day were obtained from the GISAID metadata (Elbe and Buckland-Merrett 2017; Global Initiative on Sharing All Influenza Data 2008), downloaded on 2022-03-11. Known problematic samples were removed according to the Nextstrain ‘ncov’ quality control pipeline (Hadfield et al. 2018; Nextstrain 2022). Many countries showed a pattern in which a few early cases of a focal variant (Omicron or Delta, in our analyses) were followed by many days without it, before an eventual strong rise in variant frequency. Early stochastic dynamics like this are expected, but they lead to poor fits of our deterministic model. To avoid this left tail in the data, we began the timeseries for each focal variant in each country on the first day at which three criteria were met: a variant frequency on that day of at least 10%, at least 10 cumulative variant cases, and at least 0.05% cumulative of the eventual number of variant cases. For Omicron, start dates were all in Nov and Dec 2021. For Delta, start dates ranged from Jan to Jul 2021. For each country, we ended the timeseries 9 weeks after it began for Omicron, and 15 weeks after it began for Delta; the Delta timeseries was longer because its rise was slower than Omicron’s. We further required that a country have at least 21 days with any sequence data, and at least 1000 sequences of the focal variant and of any other variants. This yielded 40 countries for Omicron and 56 countries for Delta.

To use the prevalence of Delta as a covariate for Omicron’s relative fitness, we computed for each country the proportion of variant cases that were Delta in the week prior to Omicron’s arrival. To use immunity in the population as a covariate for Omicron’s relative fitness, we computed four metrics for each country. Vaccination data were obtained from Our World in Data (OWD) (Ritchie et al. 2020), downloaded on 2022-01-06. The percent of fully vaccinated people (‘fullvax’ covariate) was defined as the number of fully vaccinated people from OWD divided by the 2020 population size, and the percent of boosted people (‘boostvax’ covariate) was defined as the number of booster shots administered divided by the population size in 2020. The vaccination levels were assumed to be constant over the study period and took the value as measured on 2022-12-15. We also considered two measures of recent infection intensity as a proxy for the level of natural immunity, ‘infected10’ and ‘infected20’, which are defined as the excess deaths per 100,000 in the 10 or 20 week period before December 1 2021 (Ritchie et al. 2020).

Finally, daily COVID-19 case counts for the selected regions and time periods were downloaded from the CSSE COVID-19 dashboard (Dong et al. 2020).

### Model

#### Renewal-equation model

Previously (van Dorp et al. 2021), we used a hierarchical logistic regression model for the increase in frequency of a new variant over time. As we noted before, this model may be sensitive to spatial and temporal variation in the effective reproduction number. This is because the region-specific effects represent the difference in growth rates of the two competing variants. This means that when the incidence is growing (*R*_*e*_ > 1), we would estimate a larger selective advantage than in an otherwise identical region where the incidence is decreasing (*R*_*e*_ < 1). The effective reproduction number within a region can also vary significantly over time, for instance due to implemented NPI in anticipation of a recently discovered variant. To account for these confounding effects, we make use of regional case count data *C*_*t*_ to estimate the time-varying effective reproduction number 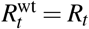 of the wild-type variant. We assume that the reproduction number of the mutant (focal variant) is given by 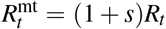. The incidence of wild-type 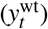 and mutant 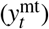 variant is governed by the discrete-time renewal equation

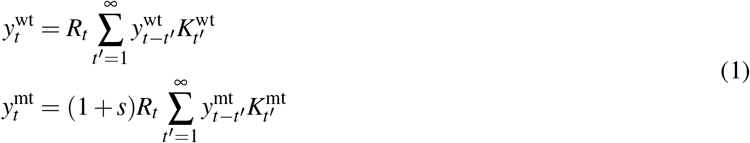

Here *K*^wt^ and *K*^mt^ denote the (discrete) probability distribution of the generation time *T*_*G*_ of wild-type and mutant variant, respectively. By allowing for *K*^wt^ ≠ *K*^mt^, we take into account that the generation time can differ between variants.

The reproduction number of the wild-type (*R*_*t*_) is modeled using a discrete-time geometric Gaussian process

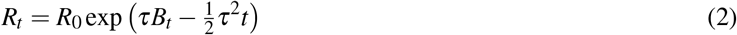

where 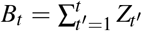 is the sum of *t* independent standard-normal increments *Z*_1_, … , *Z*_*t*_. The diffusion coefficient *τ* determines the randomness of the process and the term 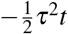 makes sure that *R*_*t*_ has constant expectation. Notice that in this notation *R*_0_ is not the basic reproduction number, but just the effective reproduction number at time *t* = 0.

To link incidence *y*_*t*_ with observed cases *C*_*t*_, we have to compute the expected number of observed cases at each observation time *t*. Let 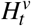 denote the probability that a person infected *t* days ago with variant *v* ∈ {wt, mt}, is counted as positive today. The number of expected cases at time *t* is then equal to

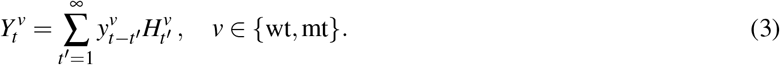

As we are primarily interested in the dynamics of the reproduction number, and not in the absolute incidence, we will assume that the total reporting probability is equal to one 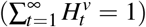. Notice that we ignore potential differences in reporting rate between the two variants. Furthermore, we make the simplifying assumption that the probability densities for reporting are identical to the probability densities for the generation time 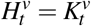, with *v* ∈ {wt, mt}).

To model the expected mutant frequency *p*_*t*_, we assume that sequences are taken from individuals that are reported positive at time *t*. Hence, we simply take

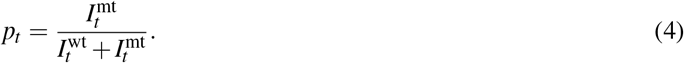

We use a beta-binomial distribution for the likelihood of the observed number of wild-type and mutant sequences. This choice allows for over-dispersion in the variant count data due to biased sampling (e.g., over-reporting of Omicron as it was emerging, Scott et al. 2021). Let 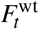 and 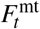 denote the number of collected sequences at time *t* for wild-type and mutant variant, respectively. We then have

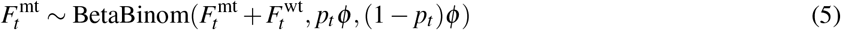

where *ϕ* determines the dispersion of the distribution. In the limit *ϕ* → ∞ this converges to a binomial distribution.

We choose a discretized Gamma distribution as generation time distribution with shape parameter *α* = 4 and mean *µ* equal to 6, 4, and 2 days for Alpha, Delta and Omicron, respectively (Hart et al. 2021; Abbott et al. 2022). The rate parameter is then given by *β* = *α/µ*. The discretization is as follows: 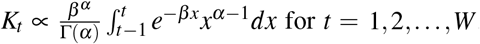, where *W* is a cutoff value equal to 15 days. The values *K*_*t*_ are then normalized such that they sum to 1.

To compute the incidence at times *t* = 1, … ,*W* , we require the incidence at times −*W* + 1, −*W* + 2, … , 0. For this, we assume that prior to time *t* = 1 the effective reproduction number was constant and equal to 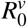. We can then compute the incidence at these prior time points by making the *ansatz* 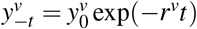 is the exponential growth rate of variant *v*. Plugging this into the renewal equation, we get

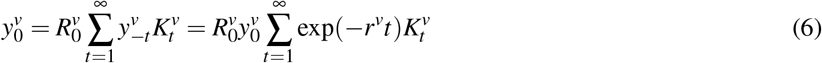

and hence 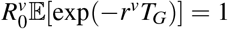, which is known as the Lotka-Euler equation. We then ignore the fact that we use a discrete-time model and use the moment-generating function of the continuous Gamma distribution, and we can find *r*^*v*^ by solving

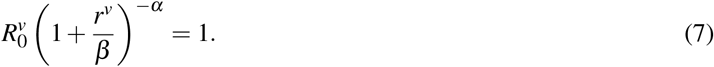

This leads to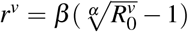. This calculation is done for both the wild type and mutant, using variant-specific parameters *α* and *β*. The initial incidence for wild-type and mutant is parameterized as follows. We introduce parameters *y*_0_ and *b*_0_, denoting the total initial incidence, and the fraction due to the mutant, respectively. Then, we set 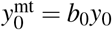 and 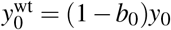.

We use a phenomenological log-normal observation model to fit the predicted case counts 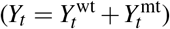 the observed case counts *C*_*t*_ ∼ Lognormal(log(*Y*_*t*_), *σ*_*C*_). To remove weekend patterns in under-reporting in the data, we aggregate the daily observations (case counts, number of sequences collected for both variants) to the week level. The same aggregation is done for the model predictions of these observations, so that we can compute the likelihood of the aggregated observations.

As we wish to estimate effects of covariates on the selective advantage in different regions, we use a Bayesian mixed-effects model (or hierarchical model) in which region-specific parameters are determined by unobserved random effects and possibly fixed effects dependent on the covariates. The structure of the model is laid out in Table 1. We focus our results on the selective advantage, *s*, of the focal variant in each country, and on the mean of its distribution across countries. We implemented the model in Stan (Stan Development Team 2019).

**Table 1:** Priors of the Bayesian mixed effects model. The matrix *M* denotes the design matrix with covariates (fullvax, boostvax, infected10, infected20). The estimated weights for *s* of these covariates are contained in the vector *w*_*s*_. The hyperparameters *µ*_*s*_ and *σ*_*s*_ determine the location and scale of the distribution of *s*. For the dispersion parameter *ϕ* , we take a prior distribution such that large values are more likely, *a priori* giving more weight to lower dispersion. *µ*_*ϕ*_ represents the mean of the dispersion parameters. For the initial incidence we took a log-normal hierarchical prior with hyper parameters *µ*_*y*0_ and *σ*_*y*0_

| parameter | prior | hyper priors |
| --- | --- | --- |
| *region-specific parameters* |  |  |
| selective advantage ($s$) | $1 + s \sim \text{Lognormal}(\mu_s + Mw_s, \sigma_s)$ | $\mu_s \sim \mathcal{N}(0, 10), \sigma_s \sim \text{Exp}(0.1)$ |
| initial mutant frequency ($b_0$) | $b_0 \sim \text{Exp}(10)$ | - |
| dispersion parameter ($\phi$) | $1/\phi \sim \text{Exp}(1/\mu_\phi)$ | $\mu_\phi \sim \text{Exp}(100)$ |
| initial incidence ($y_0$) | $y_0 \sim \text{Lognormal}(\mu_{y_0}, \sigma_{y_0})$ | $\mu_{y_0} \sim \mathcal{N}(0, 10), \sigma_{y_0} \sim \text{Exp}(0.1)$ |
| initial reproduction number ($R_0$) | $R_0 \sim \text{Lognormal}(0, 1)$ | - |
| *global parameters* |  |  |
| diffusion reproduction number ($\tau$) | $\tau \sim \text{Exp}(1)$ | - |
| standard deviation cases ($\sigma_C$) | $\sigma_C \sim \text{Exp}(1)$ | - |
| covariate weight vector ($w_s$) | $w_s \sim \mathcal{N}(0, 10)$ | - |

## Results and Discussion

Our renewal model fits provided predicted case counts, variant frequencies, and epidemic growth rates over time (Fig. 1 for selected countries, Fig. S1 for all countries). Overall the model fit the data very well, with a mean absolute error in the variant frequency of only 2.5 percentage points for data aggregated by week, or 5 percentage points for daily data (Fig. S2). Aggregating up to the week scale addressed nearly all the structural anomalies in the epidemiological data, such as non-uniform reporting by day of week, without altering the total number of cases. Even for remaining anomalous data (e.g., the spike in UK case counts due to sudden reporting of past reinfections, Fig. 1), the renewal model was able to find a reasonable estimate of the true trend.

**Figure 1:**
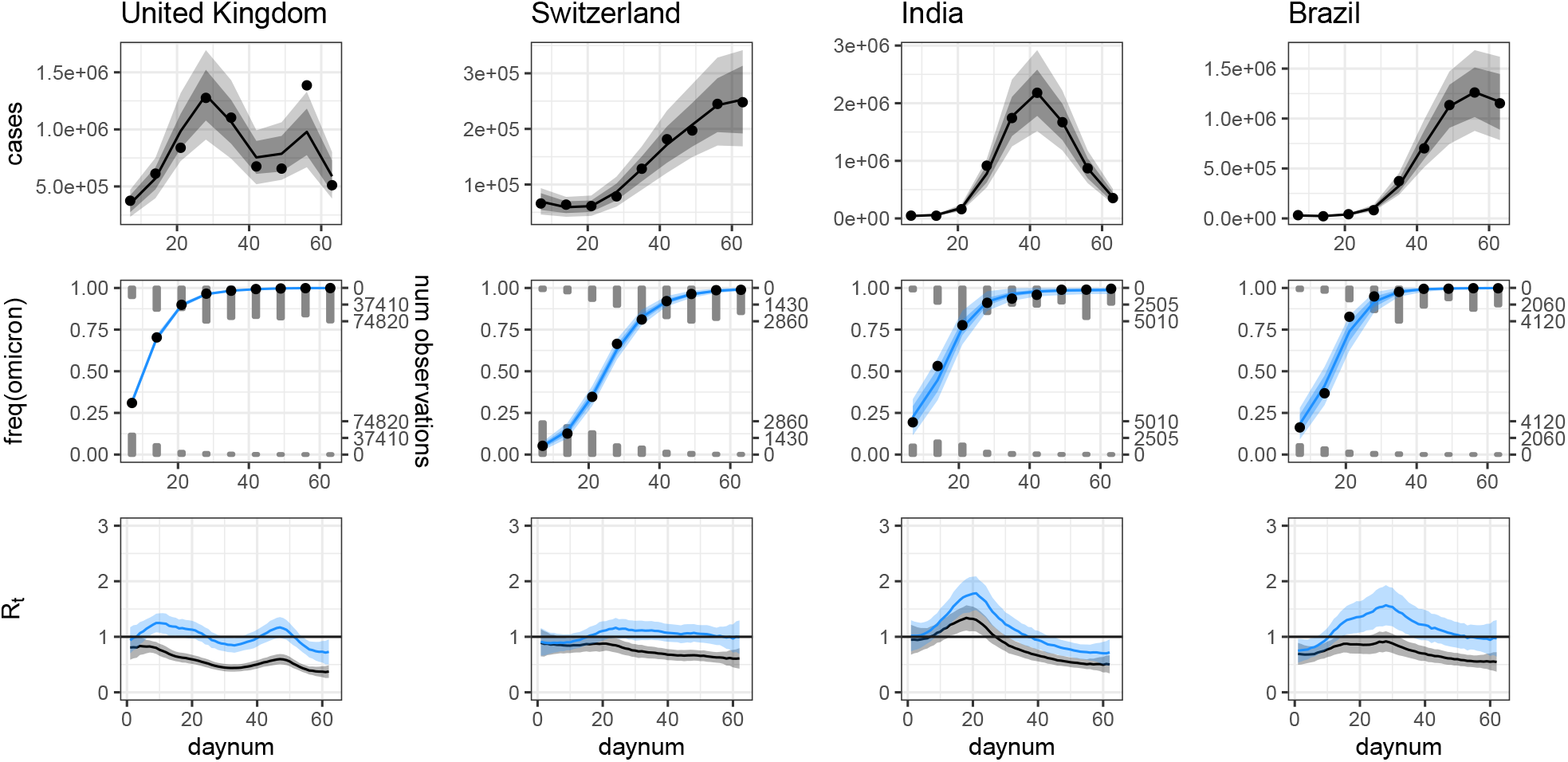
Example model fits; all countries are shown in Fig. S1. Case counts (top row) and variant counts (middle row) are binned by week, while growth rates (bottom row) are predicted by day. Model-predicted growth rates are shown for the epidemic as a whole (both variants with their true frequencies; blue) and for only the non-Omicron variant(s) (as if the Omicron frequency were zero; gray). Model predictions are shown with solid lines (median), dark shading (95% credibility interval, which includes uncertainty in the model parameters), and light shading (95% posterior predictive interval, which additionally includes sampling noise).

In almost all countries we analyzed, omicron became the dominant variant over a very short period of time, usually around 30 days. After its rise, however, Omicron was not necessarily associated with a sustained increase in the number of infections, with many countries showing a decrease in case counts after Omicron’s fixation. In countries like the United Kingdom, our model finds that nearly all of the cases in the study period could be entirely attributed to the emergence and rise of Omicron, i.e., the effective reproduction number in the wild-type circulating strains was less than one for the entire study period. In most countries, the growth rate of the wild-type variants decreases as Omicron rises in frequency, and then remains below one. This potentially reflects a combination of two factors: competition with Omicron for susceptible individuals, and the effects of interventions such as additional movement restrictions and vaccination/booster campaigns in response to Omicron’s threat. However, in India and, to a lesser extent, in Brazil the secular trend in the effective reproduction number was increase during the time that omicron was spreading. Put another way, the rise in SARS-CoV-2 during the study period in those countries cannot be explained by the spread of omicron alone and is likely due to the interaction of the increased contagiousness of Omicron and changing behavior.

### Selective advantage among countries

Comparing the model-fitted estimates of Omicron’s selective advantage, *s*, among countries shows wide variation in their values (Fig. 2A). This is consistent with previous work for other variants, which also found large heterogeneity among countries and US states (van Dorp et al. 2021; Figgins and Bedford 2021). Our model assumes that the ratio of reproduction numbers (1 + *s*) for each country is drawn from a log-normal distribution, which is simultaneously estimated from the data. This hierarchical structure minimizes the influence of structural biases in any one country’s data, leading to a more robust estimate of both the overall selection effect and the heterogeneity at country level. To examine the effect of the hierarchical model structure on our results, we also fit a non-hierarchical equivalent of our model in which *s* for each country is not affected by other countries (Fig. 2). We highlight two outcomes from this comparison: estimates for individual countries and conclusions drawn across countries.

**Figure 2:**
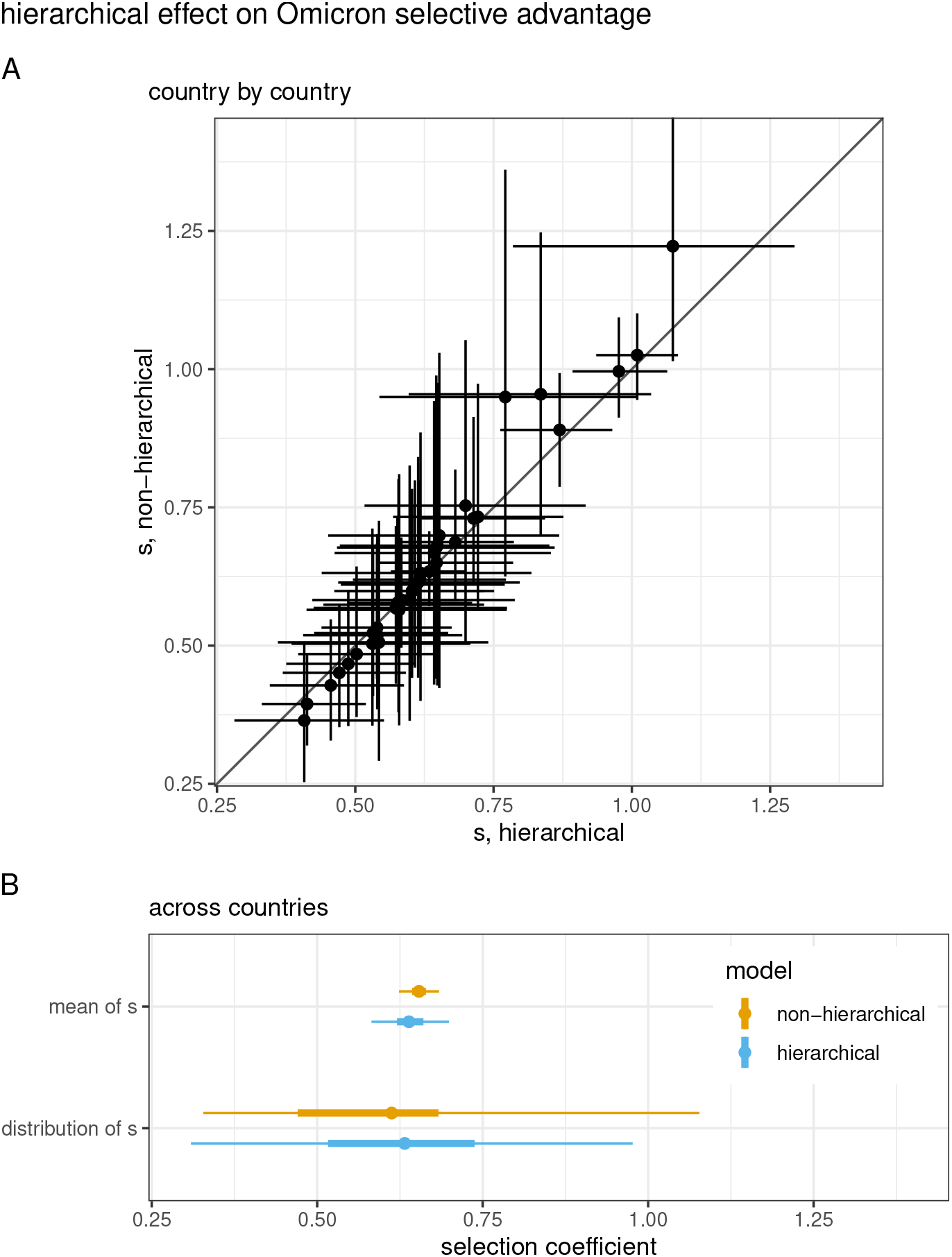
(A) Estimates of the selection coefficient, *s*, for each country, under the hierarchical or non-hierarchical models. Points and lines show the median and 95% CrI. The slope of the data is steeper than the 1:1 line, meaning that the estimates are less extreme for the hierarchical model. (B) Summaries of estimates for the mean value of *s* for Omicron, and for the distribution of values of *s* across countries, computed from the hierarchical or non-hierarchical models. Points show the median or mean, and thick and thin lines show the 50% and 95% CrI or CI.

With both the hierarchical and non-hierarchical models, we found substantial heterogeneity in the selection coefficient, *s*, at the country level for Omicron (Fig. 2A). In the hierarchical model, more extreme values of *s*—which might arise from real effects or data quality issues—are reined in by the moderating effects of other countries, although for most countries the difference in the estimate of *s* from the two methods was small. But for some countries such as Croatia, Denmark, and the UK, the non-hierarchical model substantially overestimated the selection effect. In those countries, the discrepancy is driven by the difference between the modeled and empirical frequencies of Omicron when the modeled frequencies are very high (*>*99%). Because there was often very large number of sequences collected at the end of the Omicron surge, these small discrepancies can lead to over-inflation of the estimate of *s*.

There are different ways to summarize the overall selective advantage of Omicron, encompassing the heterogeneity across countries (Fig. 2B). A first approach is to provide a typical value. For the hierarchical model this is the mean of the higher-level distribution, which has an associated uncertainty (0.64 [0.58, 0.70] 95% CrI). For the non-hierarchical model, this could be the mean of the country-level point estimates of *s*, with uncertainty summarized as the standard error of the mean (0.65 [0.62, 0.68] 95% CI). A second approach is to describe the variation across countries. For the hierarchical model this is the higher-level distribution itself, with *s* ∼ Lognormal(*µ*_*s*_, *σ*_*s*_) − 1 (Table 1), including the uncertainties on those parameters (0.64 [0.31, 0.97] 95% CrI). For the non-hierarchical model, we summarized *s* across countries by concatenating the MCMC samples from *s* for all countries (0.63 [0.33, 1.08] 95% CrI). Overall, regardless of the statistics used, Omicron has a very large selective advantage, its typical value can be estimated much more precisely than the variation among countries.

### Country-level covariates

With so much variation in *s* among countries, we asked whether causes of that variation could be identified by including country-level covariates in our model. In particular, because Omicron has been shown to evade immunity from prior infection or vaccination, we used our mixed effects model to test whether the extent of immunity in a country at the time of Omicron’s arrival explained variation in Omicron’s selective advantage. None of our immunity covariates— two measures of natural infection, and two measure of vaccination—explained a significant amount of variation in *s* among countries, however (Fig. 3). The one with the largest effect size was the proportion of people who had received full vaccination plus booster shots. We also hypothesized that Omicron might have a lesser selective advantage when it was competing primarily against Delta (which itself out-competed prior variants), but this also showed no significant effect (Fig. 3).

**Figure 3:**
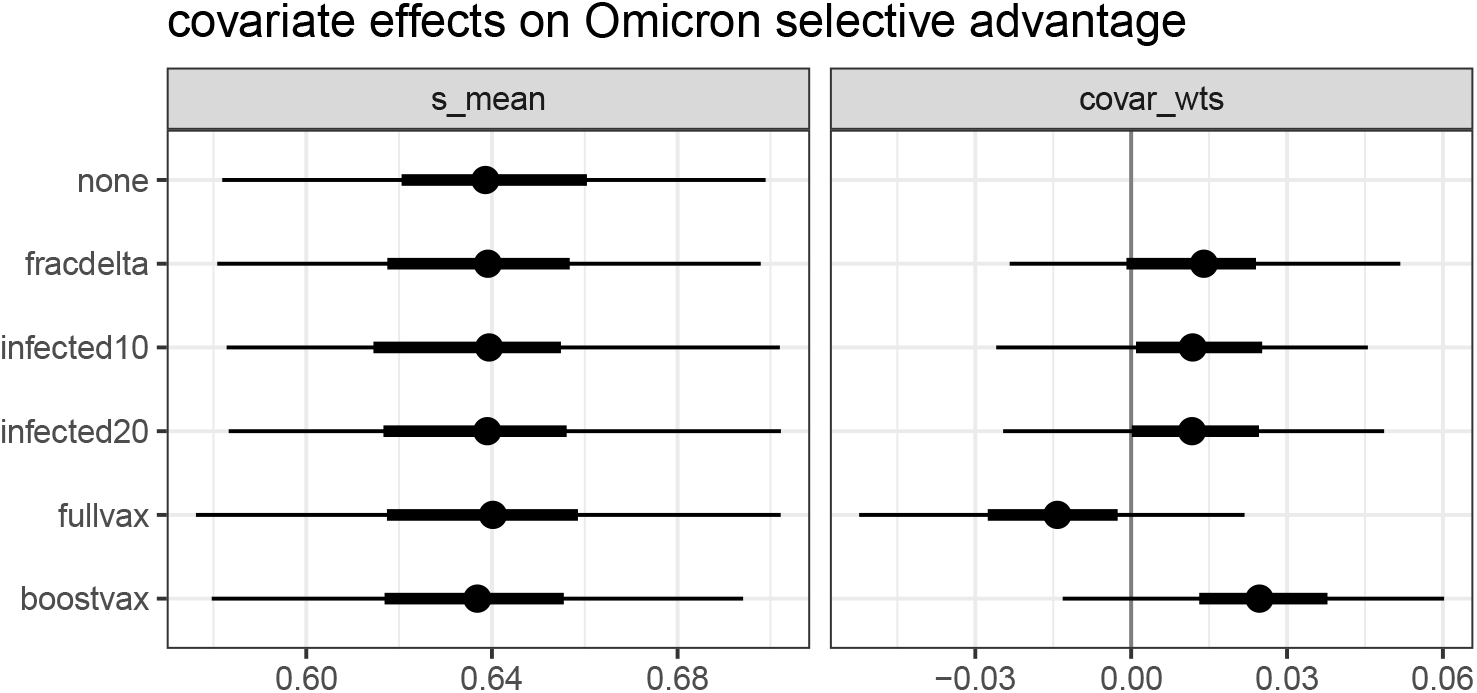
Effects of each of five country-level covariates on *s*. Plotted are the median, 50% and 95% CrI for the hierarchical mean (‘s_mean,’ essentially unchanged by including any covariate) and the estimated weight of the covariate in the model (‘covar_wts,’ all consistent with zero).

Alternatively, variation among countries could be explained not by the landscape specifically faced by Omicron, but instead by national systemic differences. With so many possible fixed country-level covariates that are likely strongly correlated, identifying specific causes is fraught. However, if any such factors do play a consistent role, we would expect their effects to be seen for variants in addition to Omicron. We therefore compared country-specific estimates of *s* for Omicron with those for Delta and found a significant correlation (Pearson’s correlation *r* = 0.3, *p* = 0.02; Fig. 4). We thus suggest that systematic differences between countries play some role in explaining the selective advantage of new variants.

**Figure 4:**
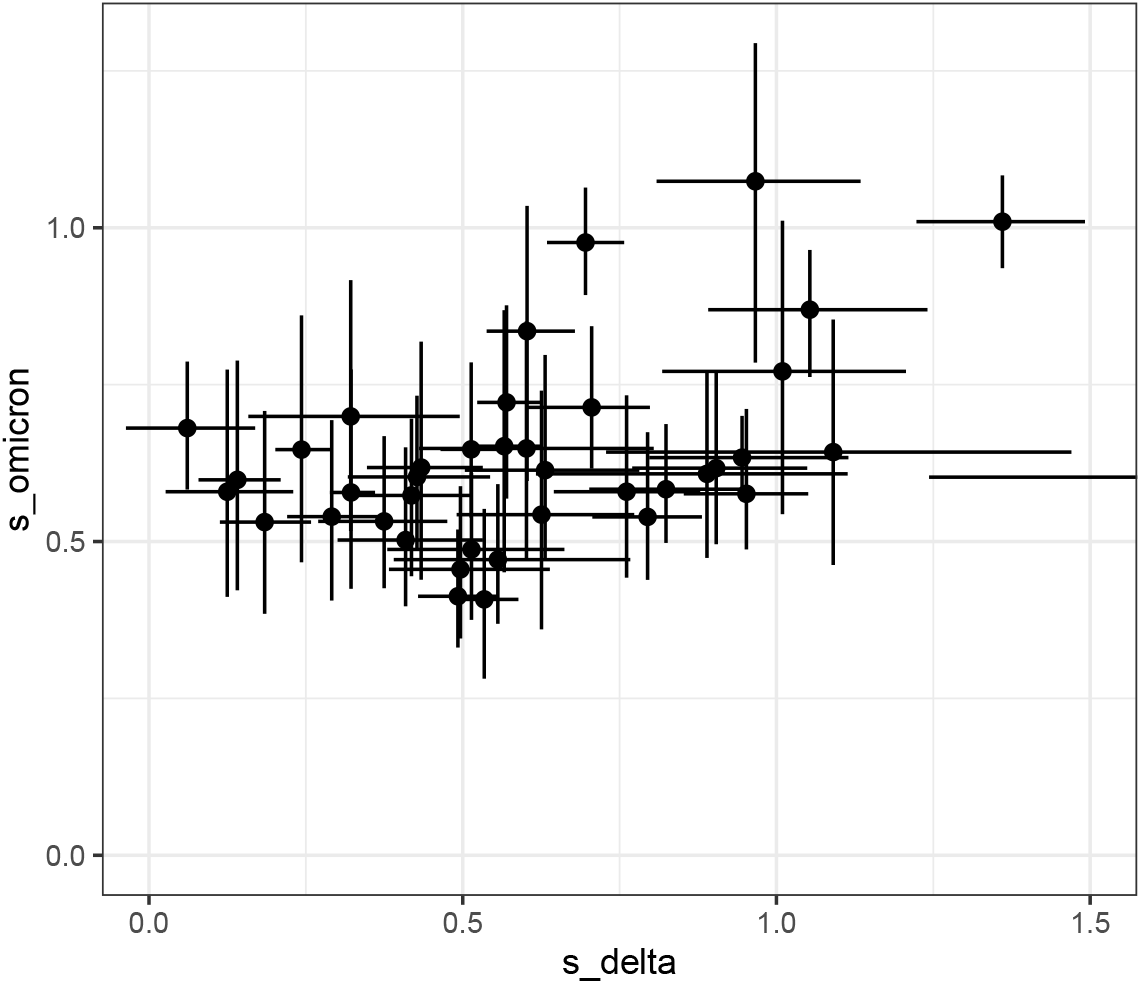
Estimates of *s* for each country, for Omicron and Delta. Plotted are the median and 95% CrI. One country not fully shown has ‘s_delta’ = 1.98.

### Conclusion

We found a very large selective advantage for Omicron (growth rate 1.6× that of the previously circulating variants, mostly Delta), but also very large heterogeneity among countries (the 95% CrI spans 1.3× to 2.0×). With selection coefficients this large, it is important to recall that their magnitude is also affected by the absolute growth rate (van Dorp et al. 2021; Chen et al. 2021). We thus used not only variant counts and a model of variant frequency dynamics, but also a renewal model that included case count data (as did Figgins and Bedford 2021).

Our hierarchical modeling approach provides a natural way to express both the overall advantage of a variant and its distribution across countries. Previous work has used a similar strategy for smaller geographic scales (Paton et al. 2022) and for mutations across lineages (Obermeyer et al. 2022). The hierarchical approach is particularly useful for global-scale analyses, in which one would like to include data from all countries even though they have vastly different data qualities and quantities.

Our mixed effects framework allows one to test hypotheses about country-level properties that might explain differences in selective advantage among countries. That such fixed properties exist is supported by the significant positive correlation between selection strength for Delta and Omicron at the country level. Identifying potential causal systemic differences may require a different approach, however, as there are many possible factors—population density, national wealth, type of governance or health care system, to name just a few— and likely strong correlations between them.

Previous work as shown by both laboratory assays (Lu et al. 2021; Cele et al. 2021; Planas et al. 2021) and population-level studies (Pearson et al. 2021; Paton et al. 2022; Pulliam et al. 2022) that Omicron better escapes natural and vaccine-induced immunity than do other variants. We thus expected to see a higher selection coefficient for Omicron in countries with more immunity, but the effects of our several country-level immunity-related covariates were not statistically significant. Our global approach has less statistical power perhaps because the covariates did not have sufficient variation among countries, or because the other as-yet-undetermined country-level factors were stronger.

Future studies of SARS-CoV-2 variant dynamics should focus on scientific explanations for heterogeneity in the spread of different variants and why some countries seem to slow the spread of new variants. The modeling framework and code presented in this paper facilitates this type of work by allowing for joint estimation of selection effects and explanatory variables.

## Data Availability

All data used in the present study are available online at http://www.gisaid.org/ https://ourworldindata.org/coronavirus and https://github.com/CSSEGISandData/COVID-19

https://github.com/eeg-lanl/sarscov2-selection

## Acknowledgements

Research presented in this article was supported by the Laboratory Directed Research and Development program of Los Alamos National Laboratory under project number 20210887ER. CHvD was funded by the National Institutes of Health (www.nih.gov), grant number U01 AI150680.

## Data availability

All data used in this study is publicly available. The scripts used for data processing and statistical inference can be found on https://github.com/eeg-lanl/sarscov2-selection

---

Figure S1: Model fits for Omicron in each country. (Provided as a separate file, omicron-fits.pdf.) Figure elements are as in Fig. 1.

**Figure S2:**
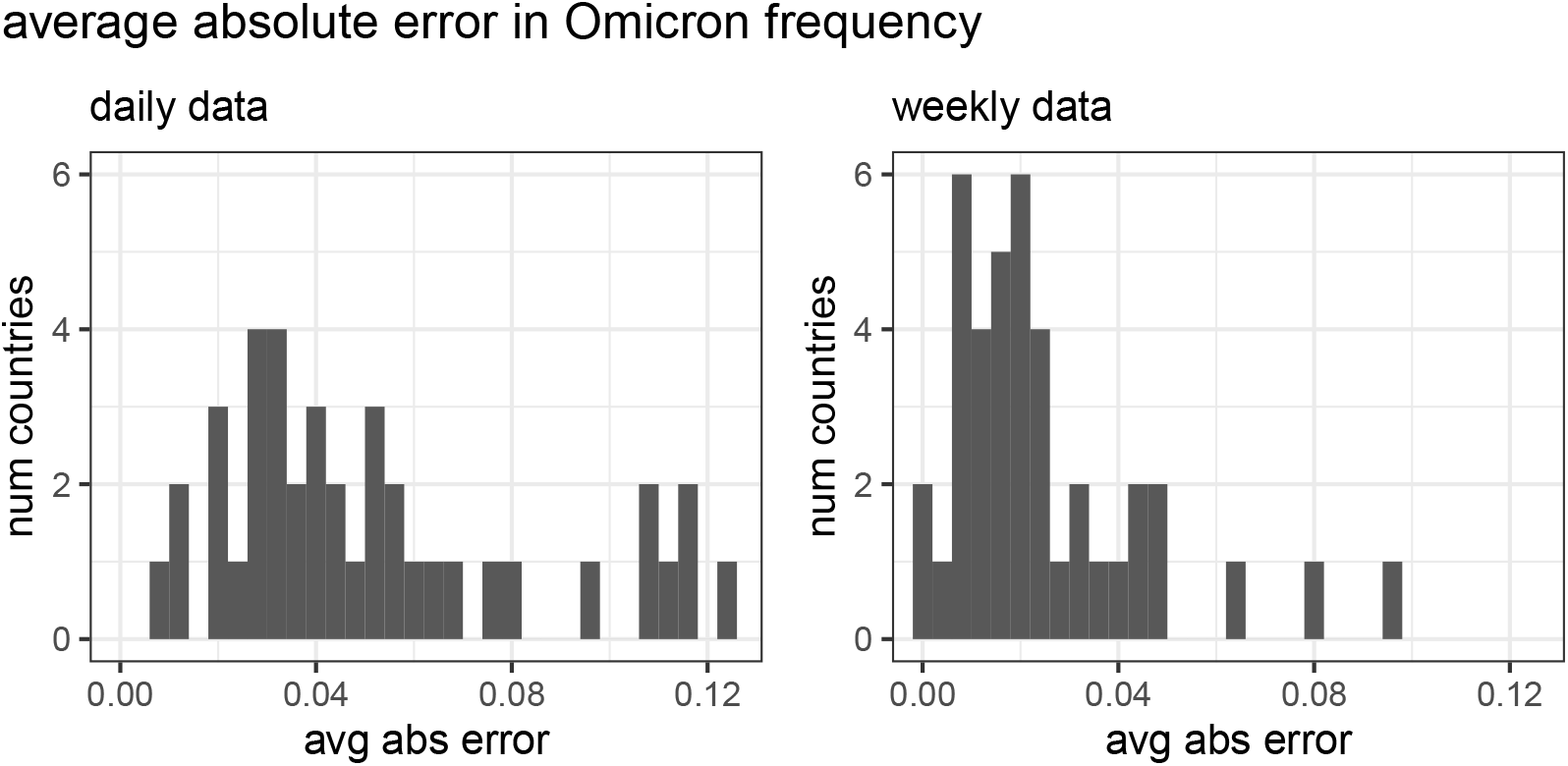
For each frequency data point (each time, each country), we computed the absolute value of its difference from the frequency predicted by our model fit. Plotted here are mean values for each country.

