## Supplementary Figure 1 for "Global estimates of the fitness advantage of SARS-CoV-2 variant Omicron"

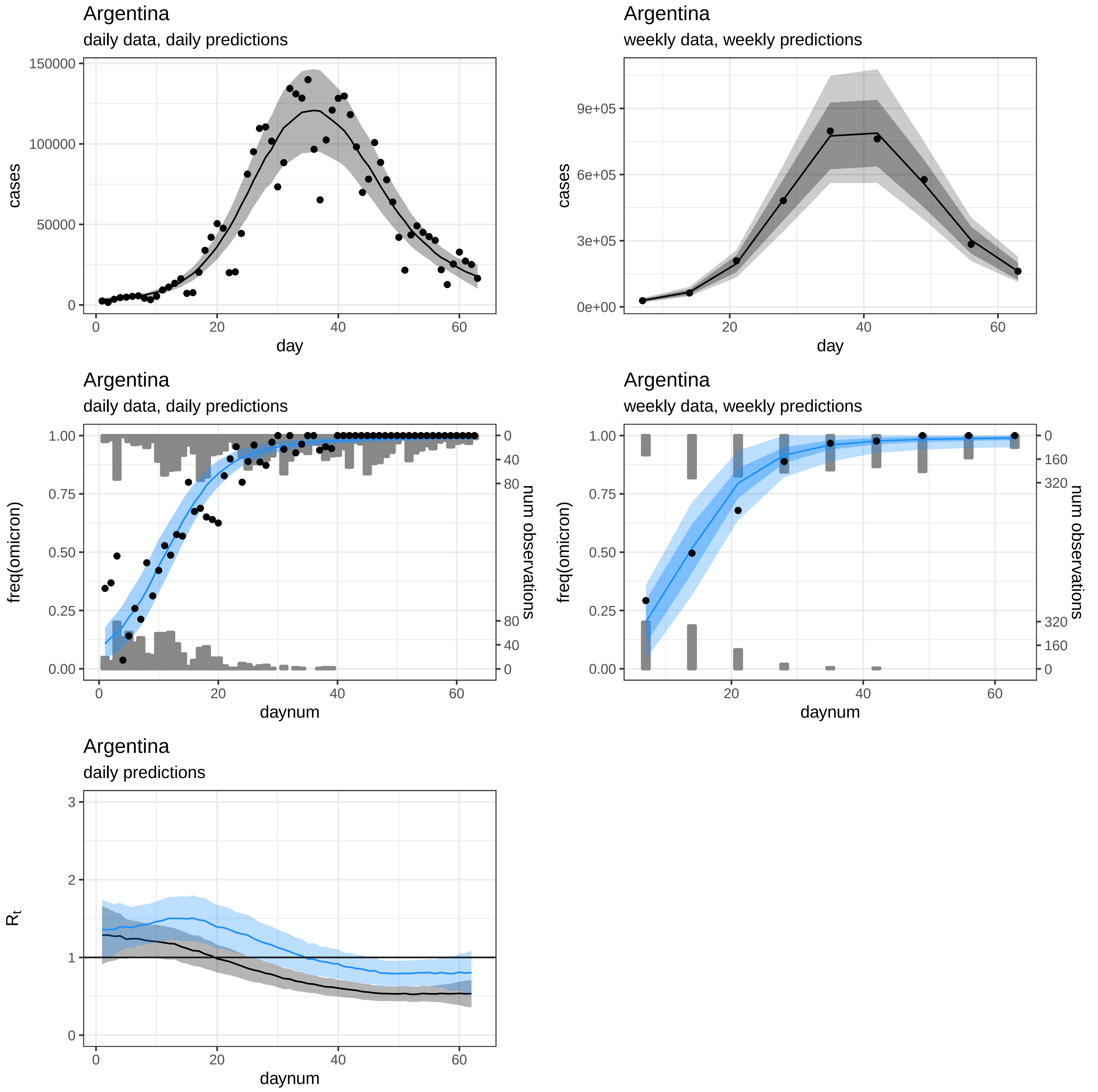
Argentina
daily data, daily predictions
cases
day
150000
100000
50000
0
0
20
40
60
Argentina
weekly data, weekly predictions
cases
day
9e+05
6e+05
3e+05
0e+00
20
40
60
Argentina
daily data, daily predictions
freq(omicron)
num observations
daynum
1.00
0.75
0.50
0.25
0.00
0
40
80
Argentina
weekly data, weekly predictions
freq(omicron)
num observations
daynum
0
160
320
Argentina
daily predictions
$R_t$
daynum
3
2
1
0

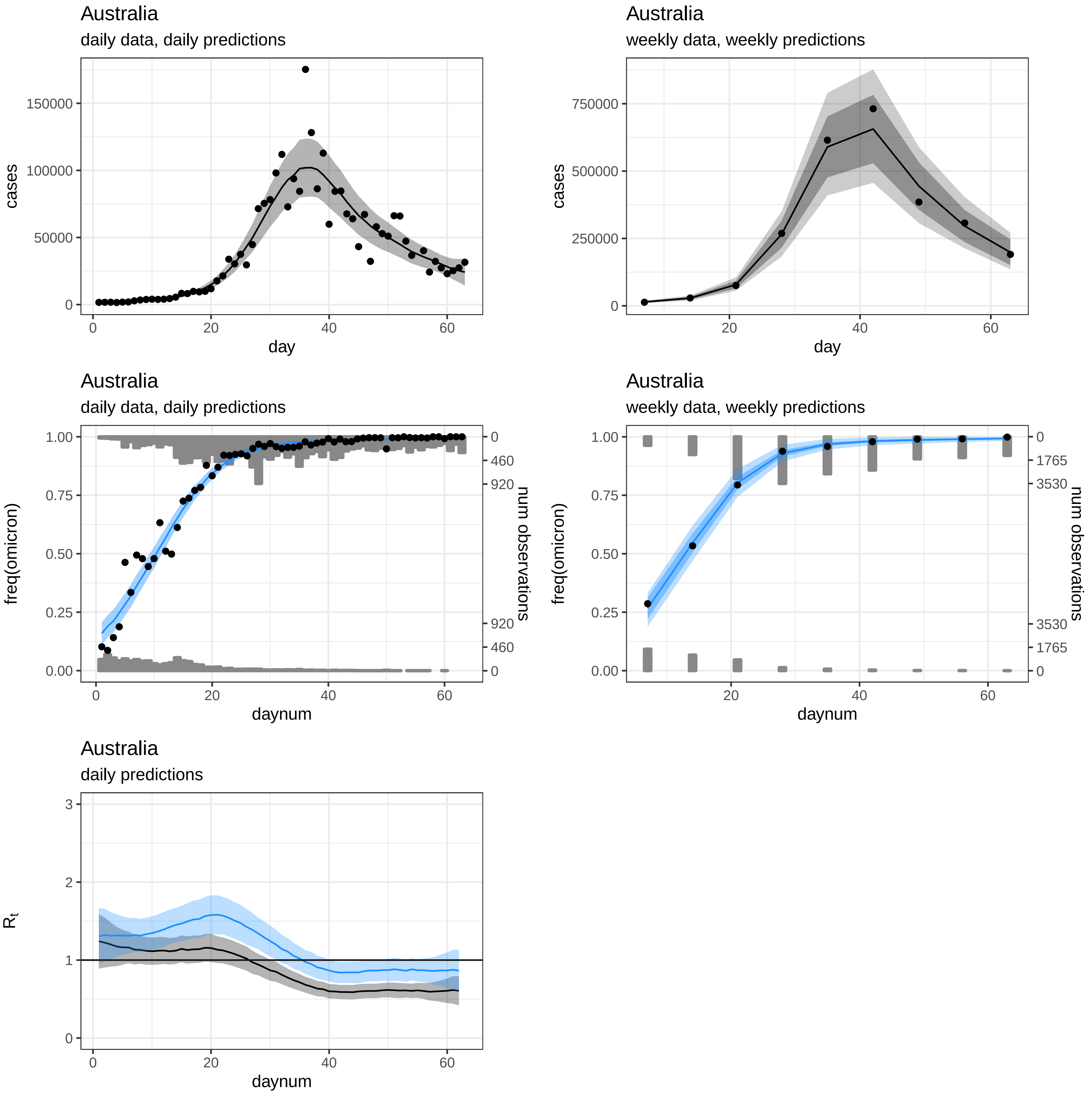
Australia
daily data, daily predictions
cases
150000
100000
50000
0
0
20
40
60
day
Australia
weekly data, weekly predictions
cases
750000
500000
250000
0
20
40
60
day
Australia
daily data, daily predictions
freq(omicron)
1.00
0.75
0.50
0.25
0.00
0
460
920
920
460
0
num observations
0
20
40
60
daynum
Australia
weekly data, weekly predictions
freq(omicron)
1.00
0.75
0.50
0.25
0.00
0
1765
3530
3530
1765
0
num observations
20
40
60
daynum
Australia
daily predictions
$R_t$
3
2
1
0
0
20
40
60
daynum

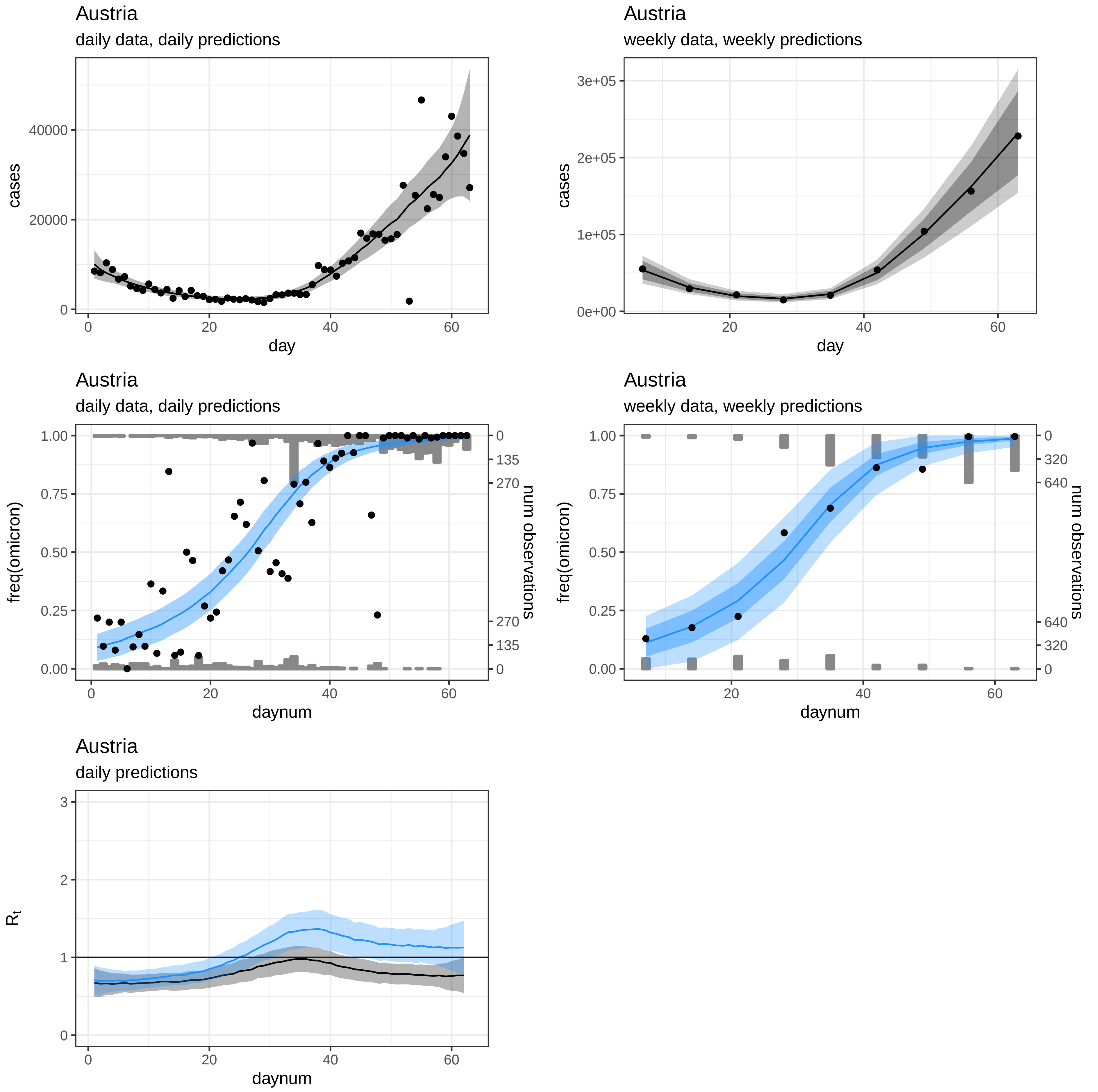
Austria
daily data, daily predictions
cases
day
Austria
weekly data, weekly predictions
cases
day
Austria
daily data, daily predictions
freq(omicron)
num observations
daynum
Austria
weekly data, weekly predictions
freq(omicron)
num observations
daynum
Austria
daily predictions
$R_t$
daynum

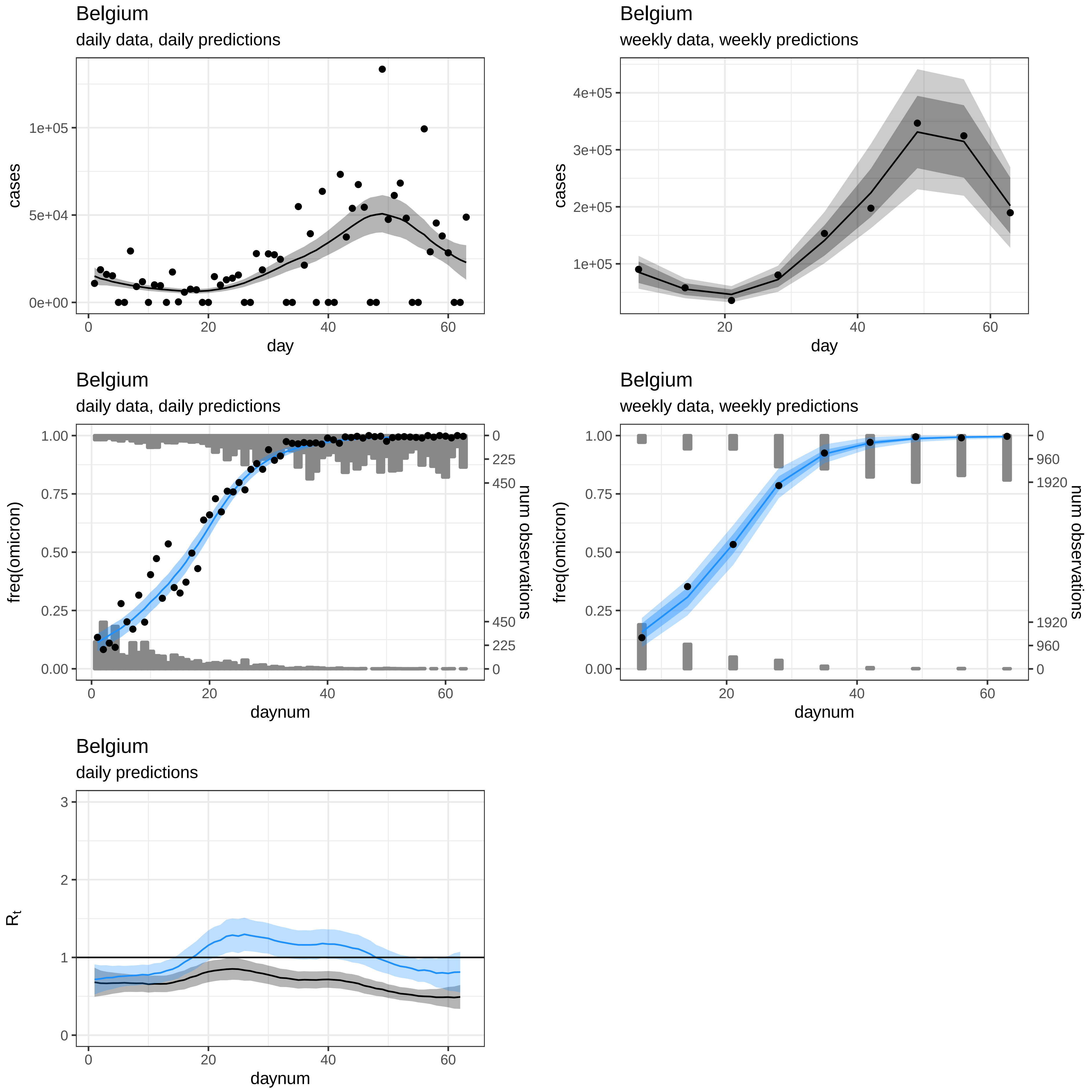
Belgium
daily data, daily predictions
cases
1e+05
5e+04
0e+00
0
20
40
60
day
Belgium
weekly data, weekly predictions
cases
4e+05
3e+05
2e+05
1e+05
20
40
60
day
Belgium
daily data, daily predictions
freq(omicron)
1.00
0.75
0.50
0.25
0.00
num observations
0
225
450
450
225
0
0
20
40
60
daynum
Belgium
weekly data, weekly predictions
freq(omicron)
1.00
0.75
0.50
0.25
0.00
num observations
0
960
1920
1920
960
0
20
40
60
daynum
Belgium
daily predictions
$R_t$
3
2
1
0
0
20
40
60
daynum

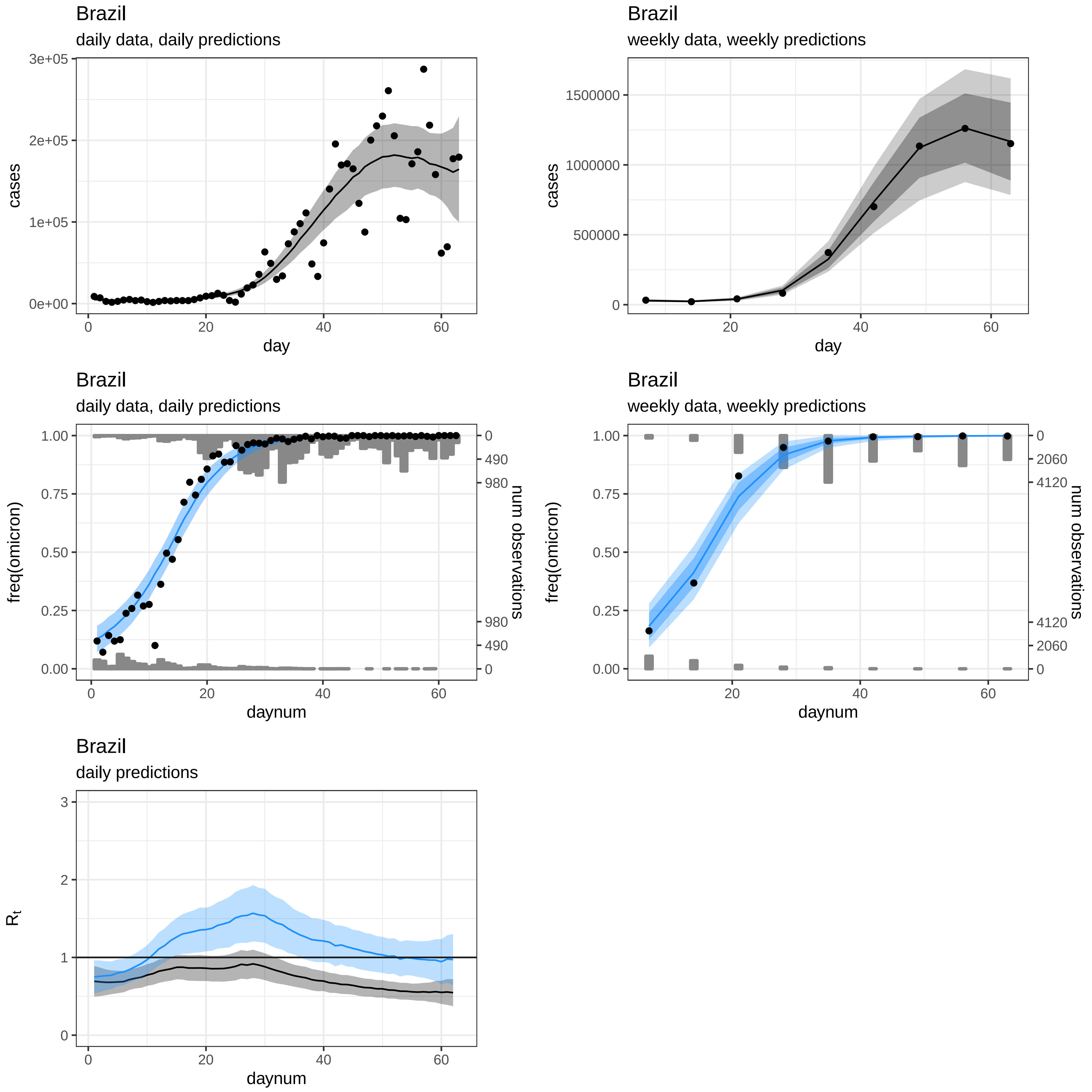
Brazil
daily data, daily predictions
cases
3e+05
2e+05
1e+05
0e+00
0
20
40
60
day
Brazil
weekly data, weekly predictions
cases
1500000
1000000
500000
0
20
40
60
day
Brazil
daily data, daily predictions
freq(omicron)
1.00
0.75
0.50
0.25
0.00
num observations
0
490
980
980
490
0
0
20
40
60
daynum
Brazil
weekly data, weekly predictions
freq(omicron)
1.00
0.75
0.50
0.25
0.00
num observations
0
2060
4120
4120
2060
0
20
40
60
daynum
Brazil
daily predictions
$R_t$
3
2
1
0
0
20
40
60
daynum

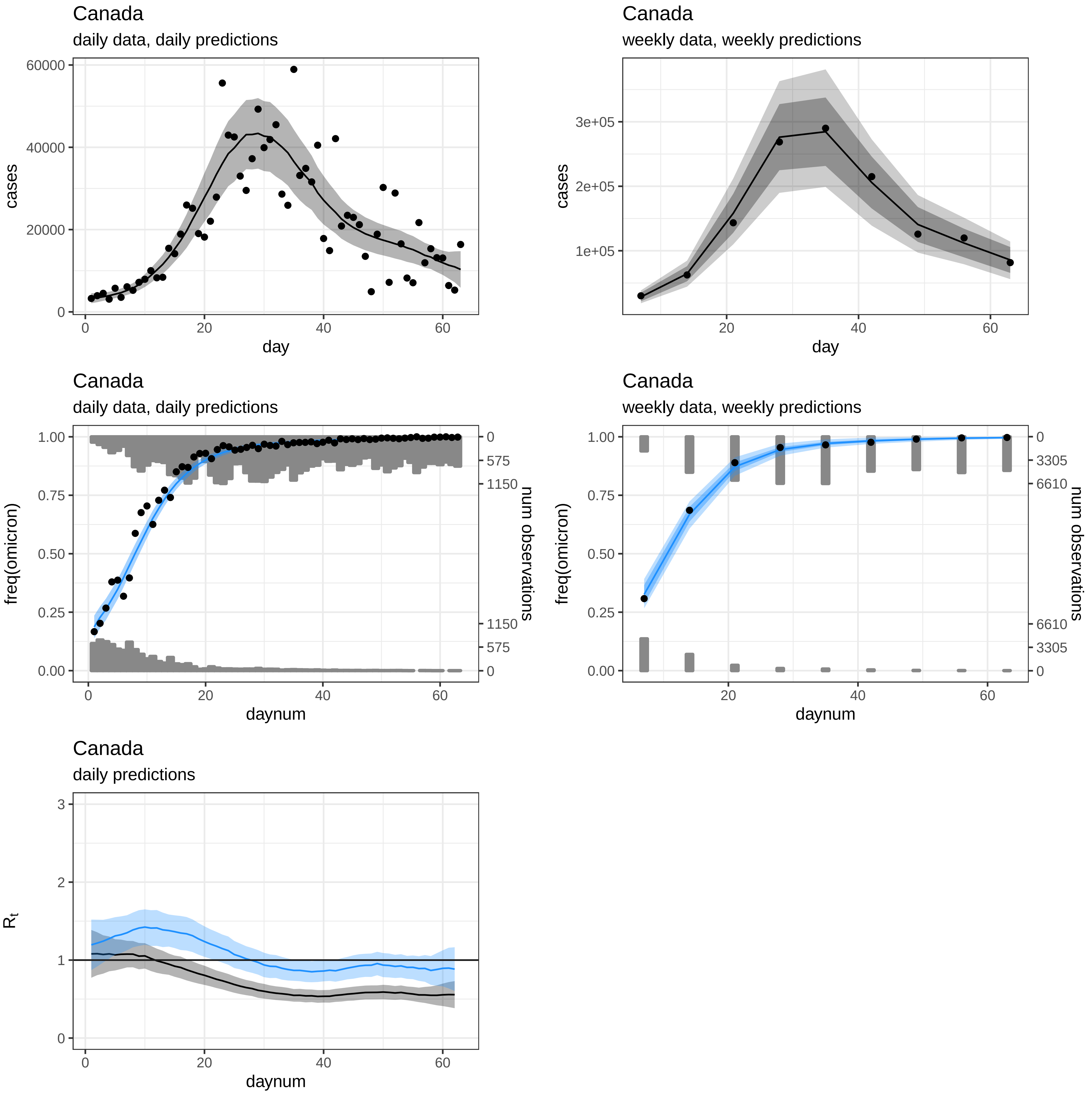
Canada
daily data, daily predictions
cases
day
Canada
weekly data, weekly predictions
cases
day
Canada
daily data, daily predictions
freq(omicron)
num observations
daynum
Canada
weekly data, weekly predictions
freq(omicron)
num observations
daynum
Canada
daily predictions
$R_t$
daynum

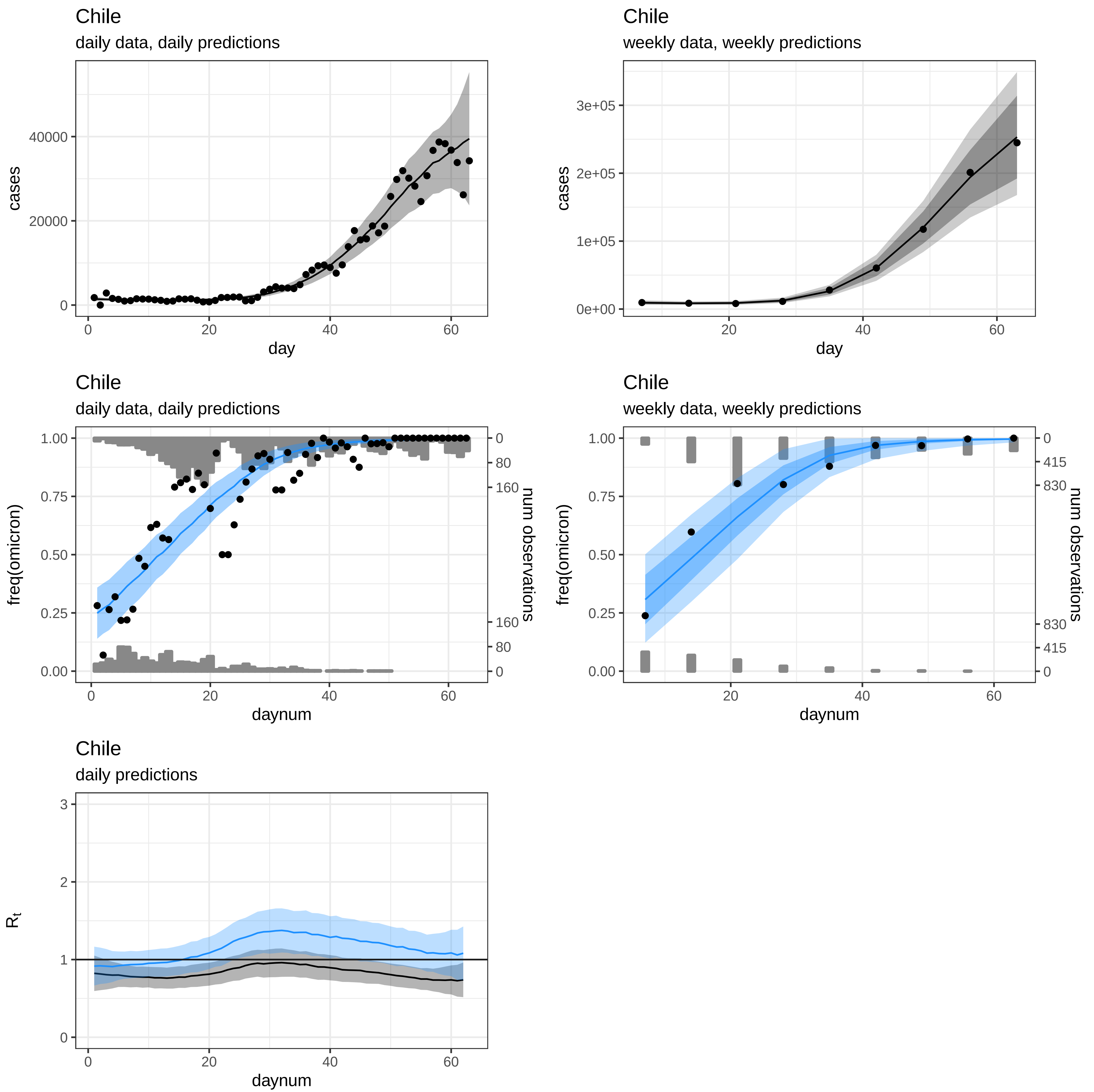
Chile
daily data, daily predictions
cases
day
Chile
weekly data, weekly predictions
cases
day
Chile
daily data, daily predictions
freq(omicron)
num observations
daynum
Chile
weekly data, weekly predictions
freq(omicron)
num observations
daynum
Chile
daily predictions
$R_t$
daynum

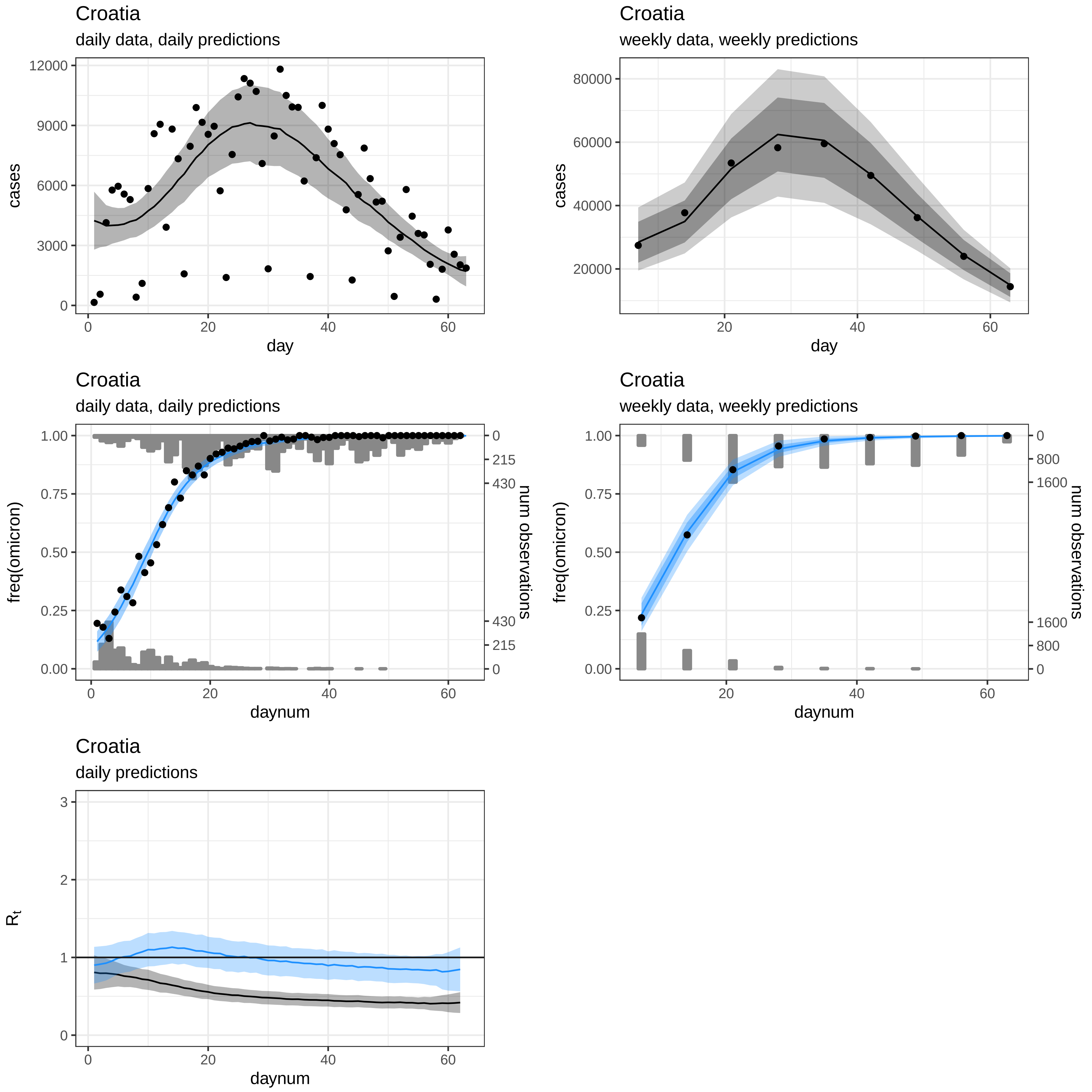

Croatia
daily data, daily predictions
cases
day
12000
9000
6000
3000
0
0
20
40
60
Croatia
weekly data, weekly predictions
cases
day
80000
60000
40000
20000
20
40
60
Croatia
daily data, daily predictions
freq(omicron)
num observations
daynum
1.00
0.75
0.50
0.25
0.00
0
215
430
430
215
0
0
20
40
60
Croatia
weekly data, weekly predictions
freq(omicron)
num observations
daynum
1.00
0.75
0.50
0.25
0.00
0
800
1600
1600
800
0
20
40
60
Croatia
daily predictions
$R_t$
daynum
3
2
1
0
0
20
40
60

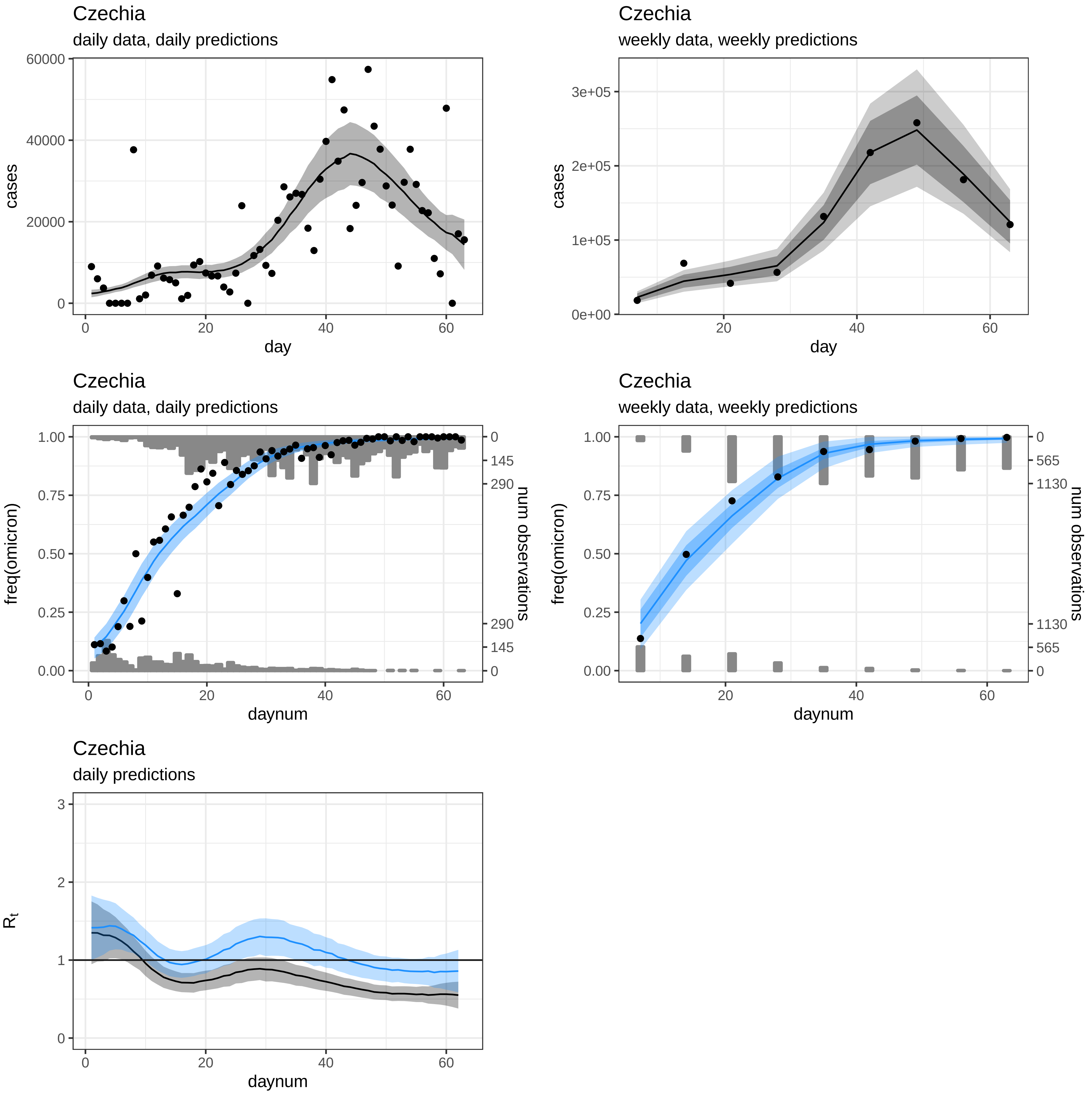
Czechia
daily data, daily predictions
cases
60000
40000
20000
0
0
20
40
60
day
Czechia
weekly data, weekly predictions
cases
3e+05
2e+05
1e+05
0e+00
20
40
60
day
Czechia
daily data, daily predictions
freq(omicron)
1.00
0.75
0.50
0.25
0.00
num observations
0
145
290
290
145
0
0
20
40
60
daynum
Czechia
weekly data, weekly predictions
freq(omicron)
1.00
0.75
0.50
0.25
0.00
num observations
0
565
1130
1130
565
0
20
40
60
daynum
Czechia
daily predictions
$R_t$
3
2
1
0
0
20
40
60
daynum

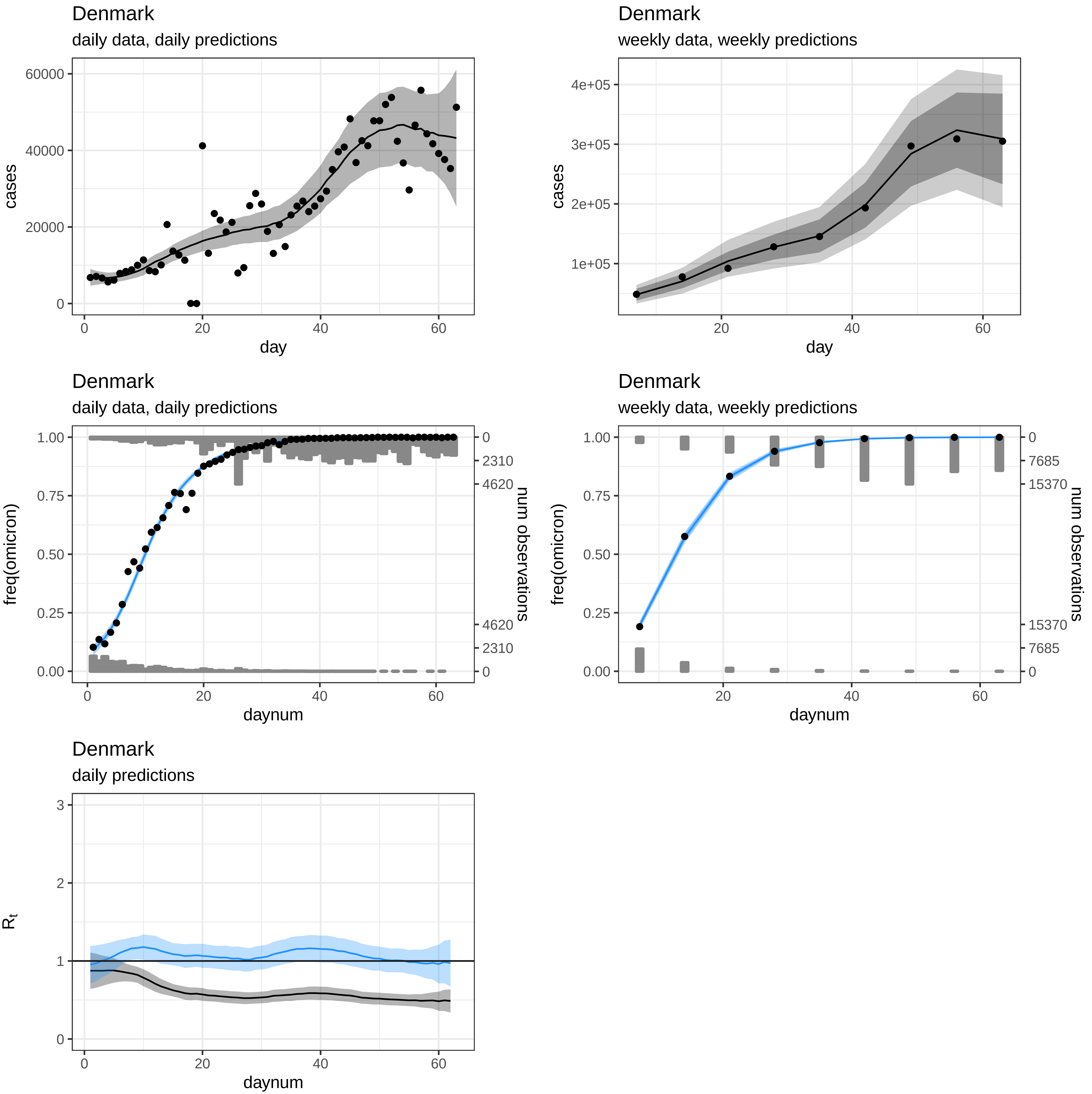
Denmark
daily data, daily predictions
cases
day
60000
40000
20000
0
20
40
60
Denmark
weekly data, weekly predictions
4e+05
3e+05
2e+05
1e+05
Denmark
daily data, daily predictions
freq(omicron)
num observations
daynum
1.00
0.75
0.50
0.25
0.00
2310
4620
Denmark
weekly data, weekly predictions
7685
15370
Denmark
daily predictions
$R_t$
3
2
1

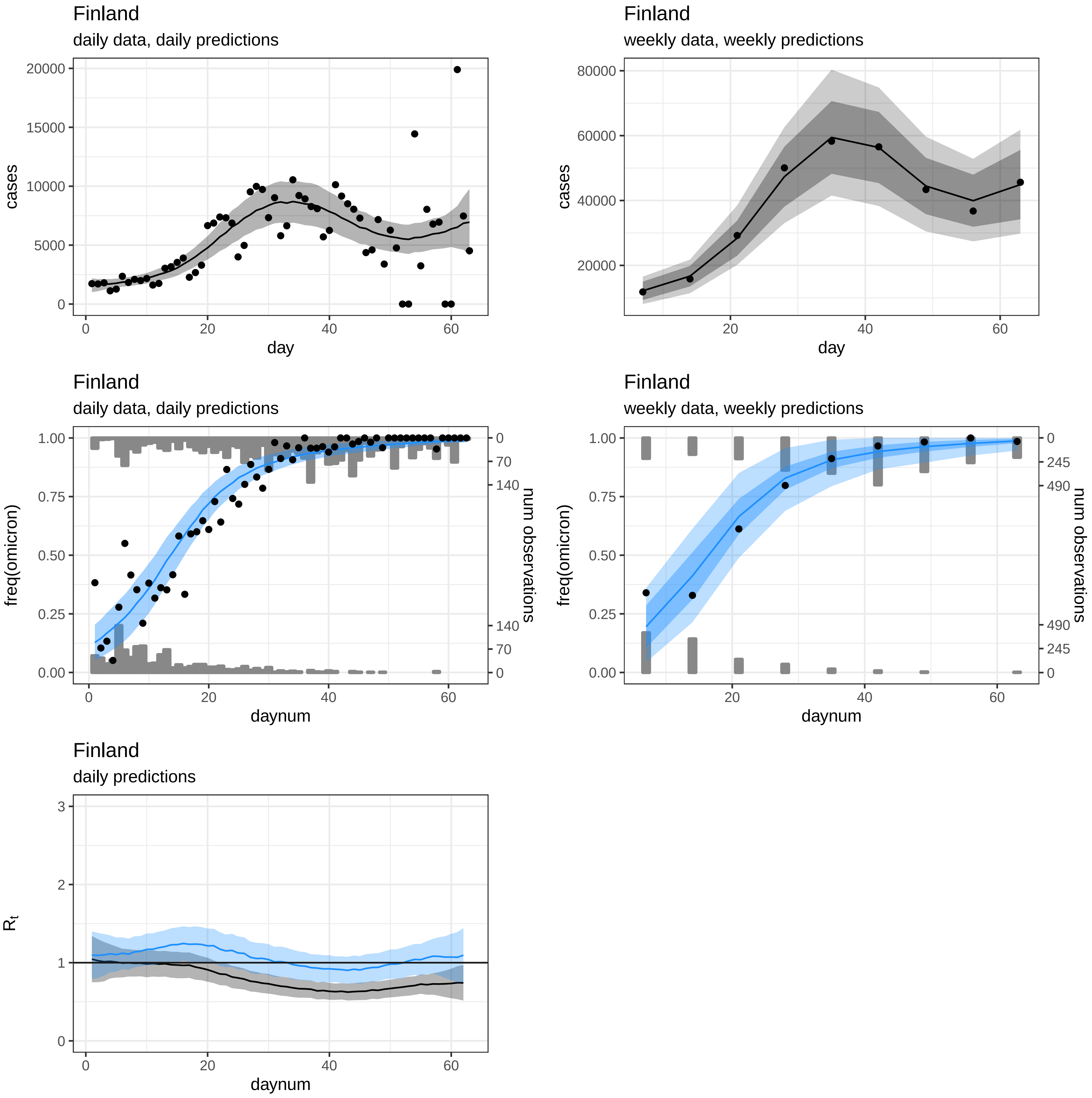
Finland
daily data, daily predictions
cases
day
Finland
weekly data, weekly predictions
cases
day
Finland
daily data, daily predictions
freq(omicron)
num observations
daynum
Finland
weekly data, weekly predictions
freq(omicron)
num observations
daynum
Finland
daily predictions
$R_t$
daynum

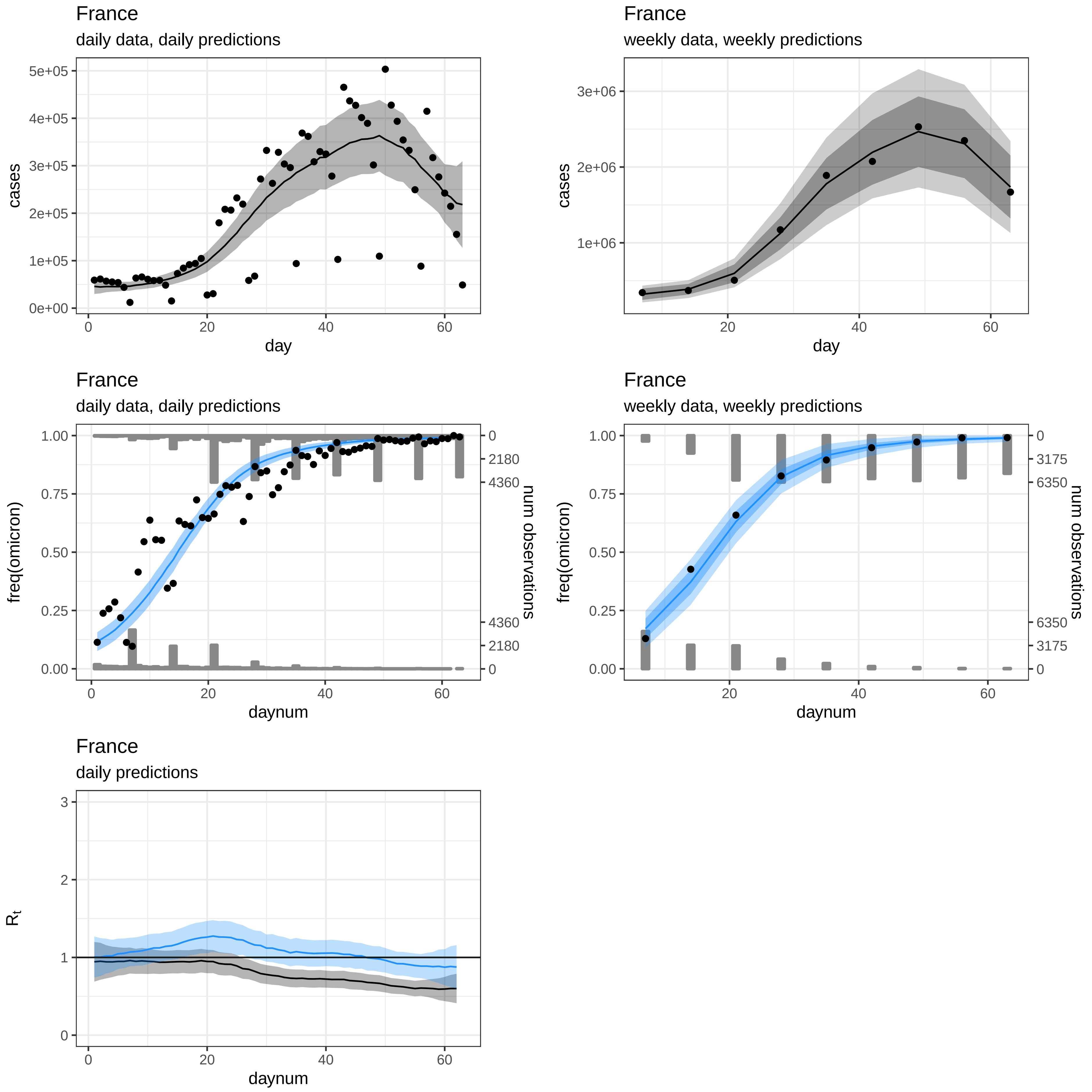
France
daily data, daily predictions
cases
day
France
weekly data, weekly predictions
cases
day
France
daily data, daily predictions
freq(omicron)
num observations
daynum
France
weekly data, weekly predictions
freq(omicron)
num observations
daynum
France
daily predictions
$R_t$
daynum

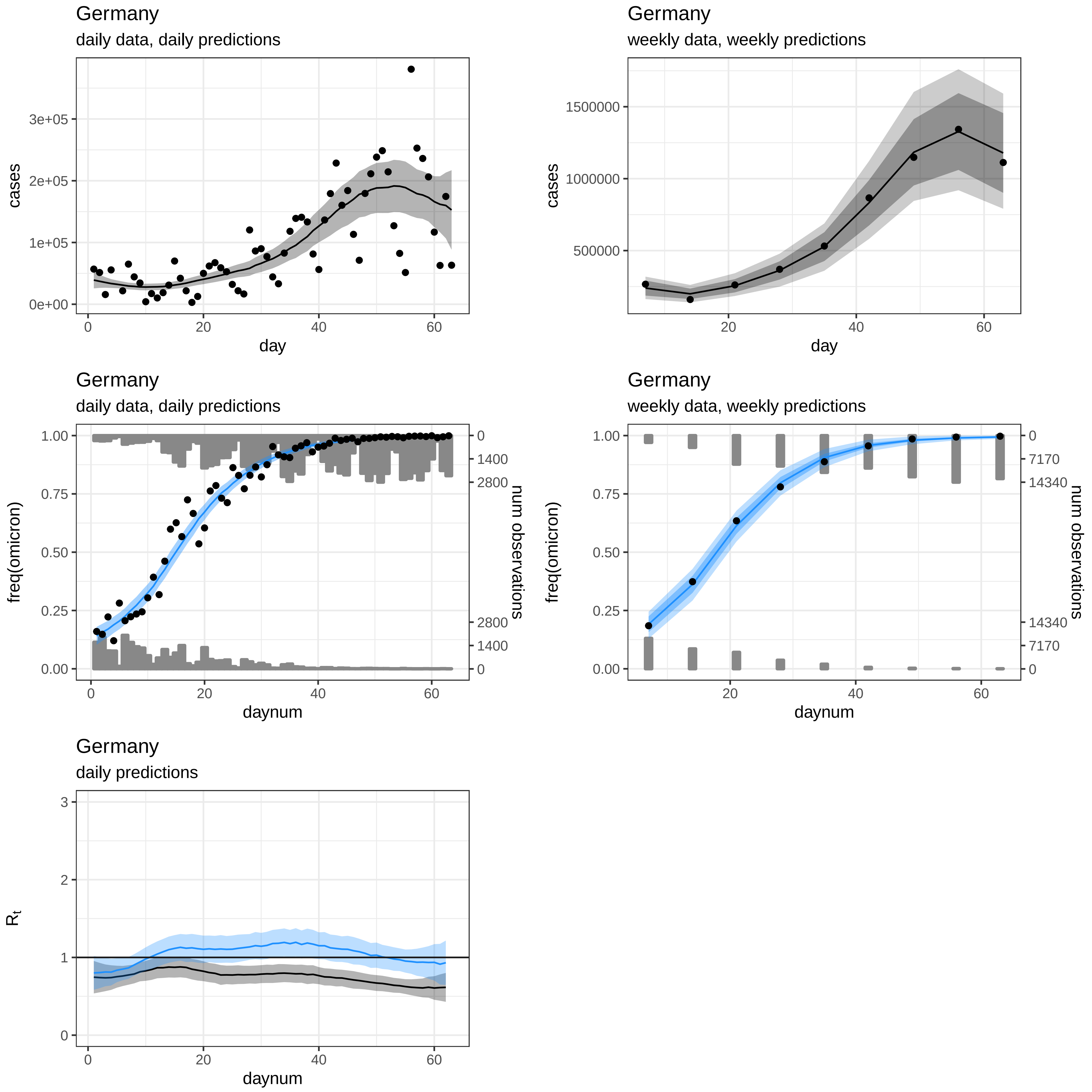
Germany
daily data, daily predictions
cases
day
3e+05
2e+05
1e+05
0e+00
0
20
40
60
Germany
weekly data, weekly predictions
cases
day
1500000
1000000
500000
20
40
60
Germany
daily data, daily predictions
freq(omicron)
num observations
daynum
1.00
0.75
0.50
0.25
0.00
0
1400
2800
2800
1400
0
Germany
weekly data, weekly predictions
freq(omicron)
num observations
daynum
0
7170
14340
14340
7170
0
Germany
daily predictions
$R_t$
daynum
3
2
1
0

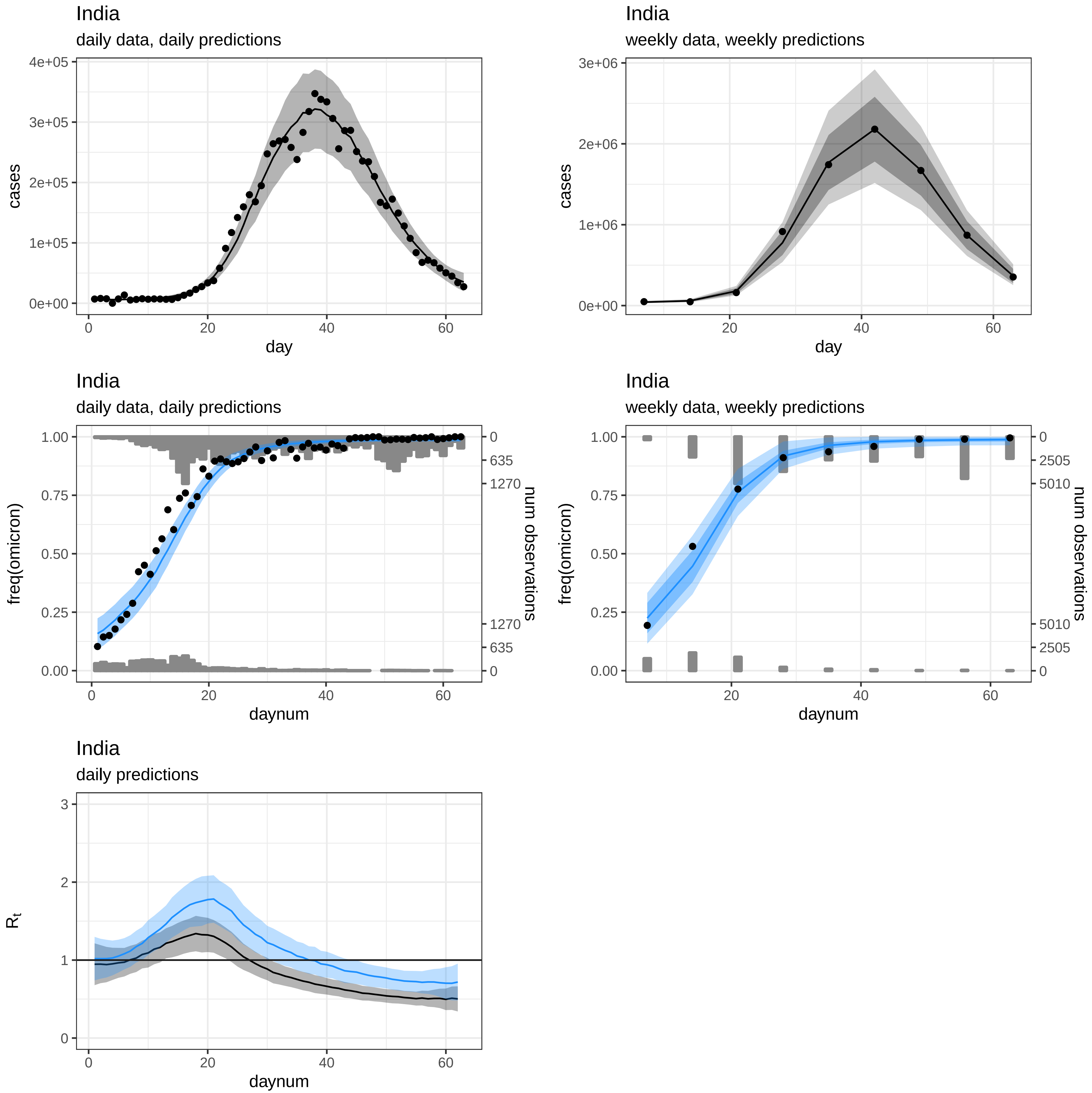
India
daily data, daily predictions
cases
day
India
weekly data, weekly predictions
cases
day
India
daily data, daily predictions
freq(omicron)
num observations
daynum
India
weekly data, weekly predictions
freq(omicron)
num observations
daynum
India
daily predictions
$R_t$
daynum

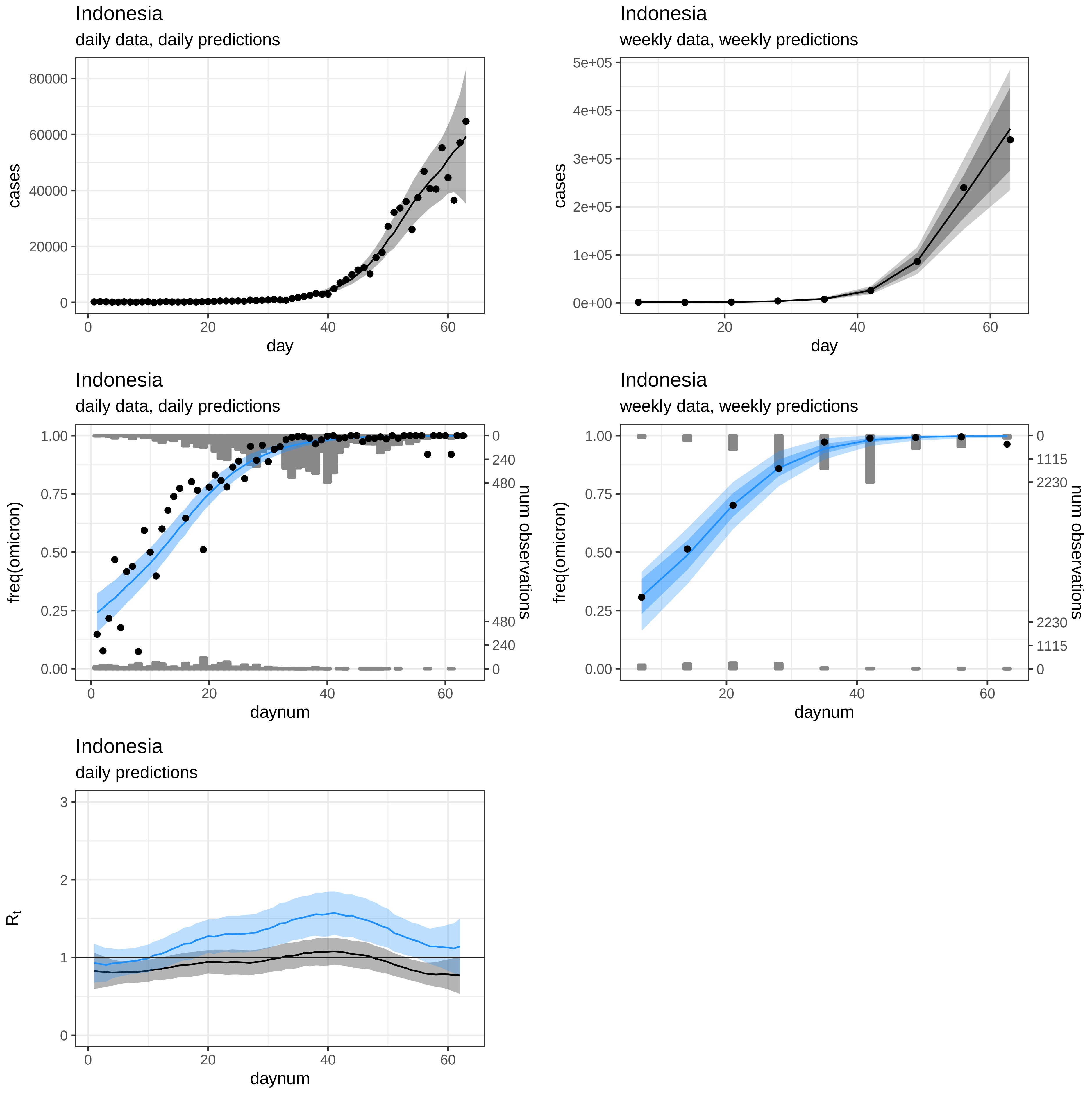
Indonesia
daily data, daily predictions
cases
day
Indonesia
weekly data, weekly predictions
cases
day
Indonesia
daily data, daily predictions
freq(omicron)
num observations
daynum
Indonesia
weekly data, weekly predictions
freq(omicron)
num observations
daynum
Indonesia
daily predictions
$R_t$
daynum

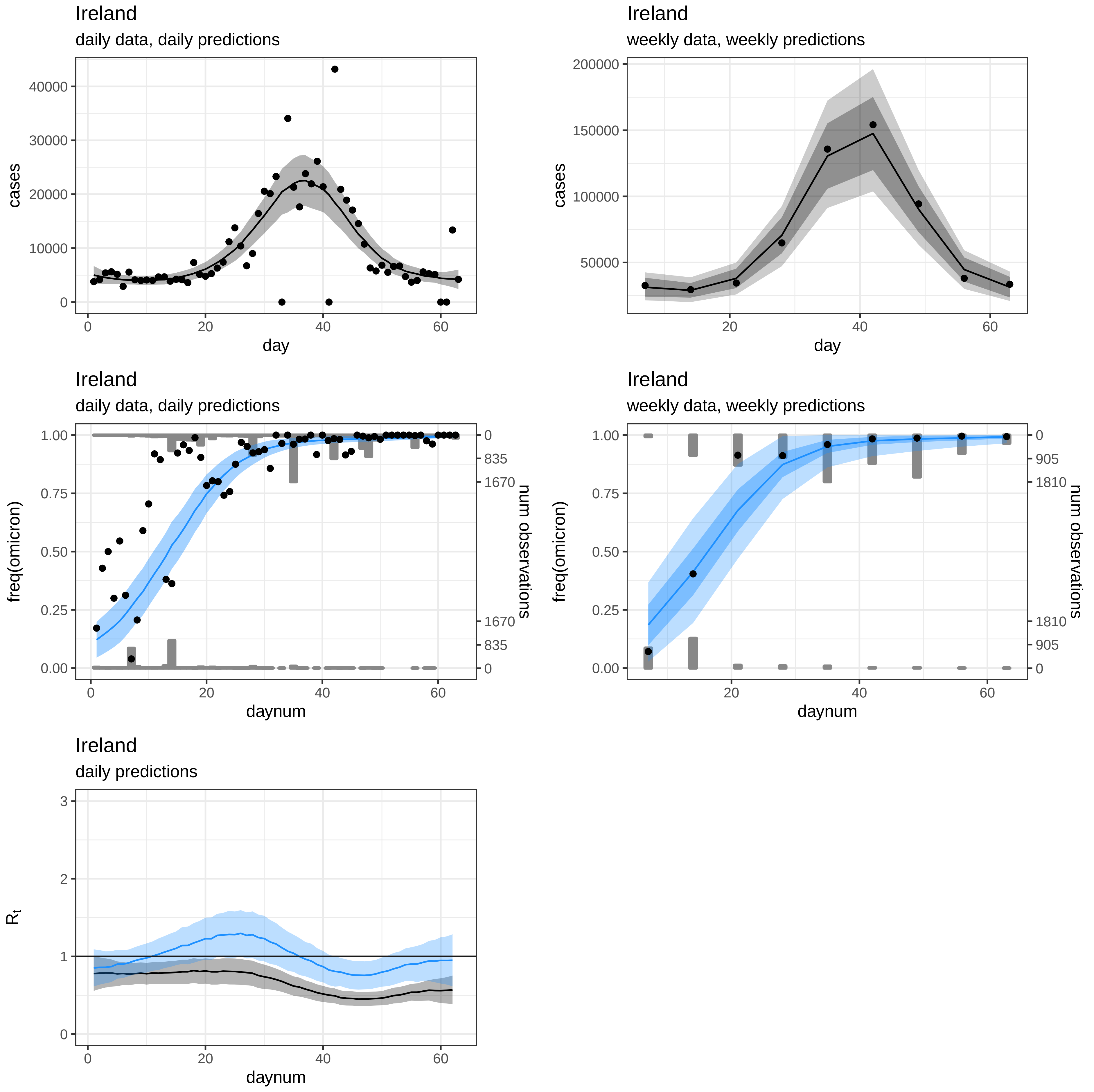
Ireland
daily data, daily predictions
cases
day
Ireland
weekly data, weekly predictions
cases
day
Ireland
daily data, daily predictions
freq(omicron)
num observations
daynum
Ireland
weekly data, weekly predictions
freq(omicron)
num observations
daynum
Ireland
daily predictions
$R_t$
daynum

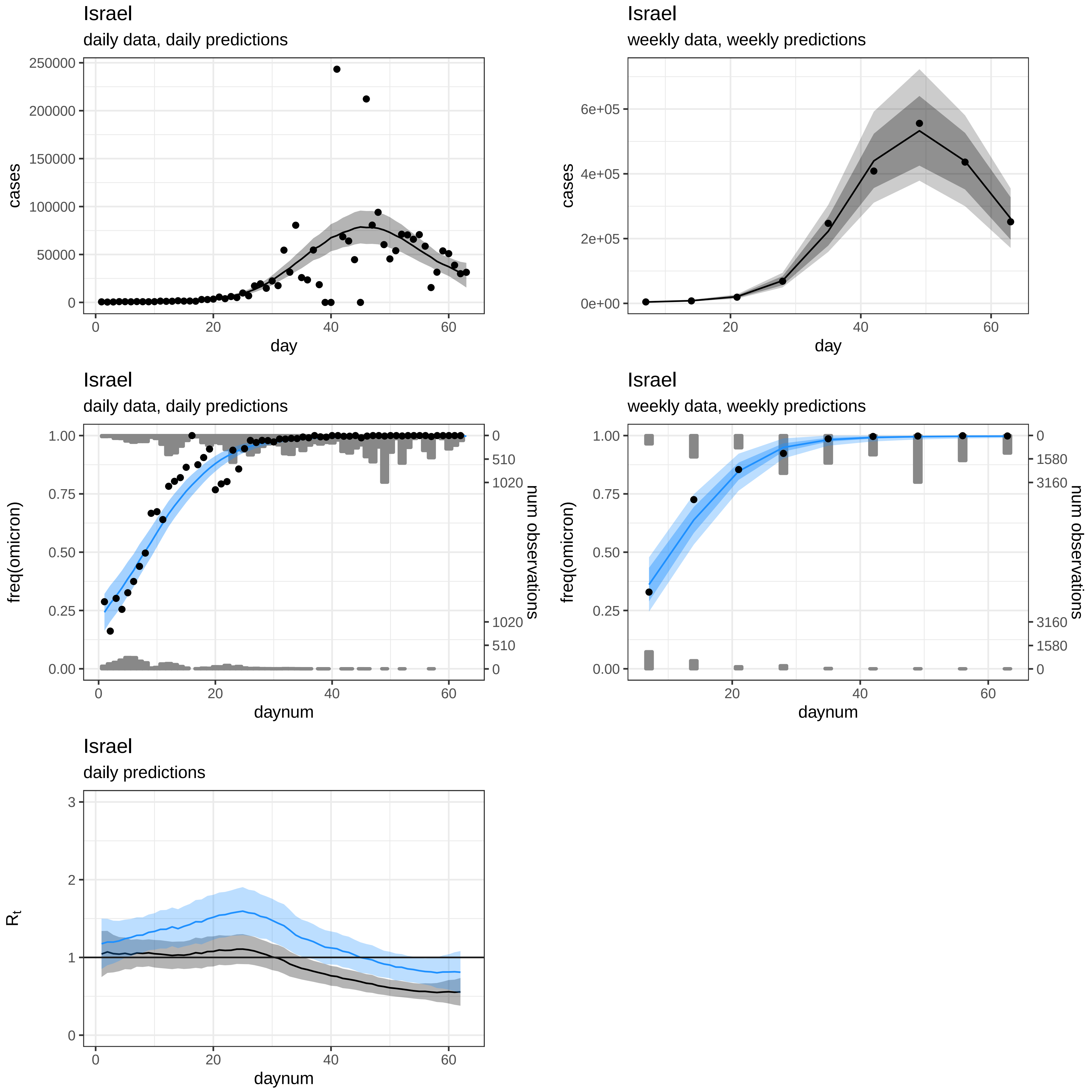
Israel
daily data, daily predictions
cases
day
Israel
weekly data, weekly predictions
cases
day
Israel
daily data, daily predictions
freq(omicron)
num observations
daynum
Israel
weekly data, weekly predictions
freq(omicron)
num observations
daynum
Israel
daily predictions
$R_t$
daynum

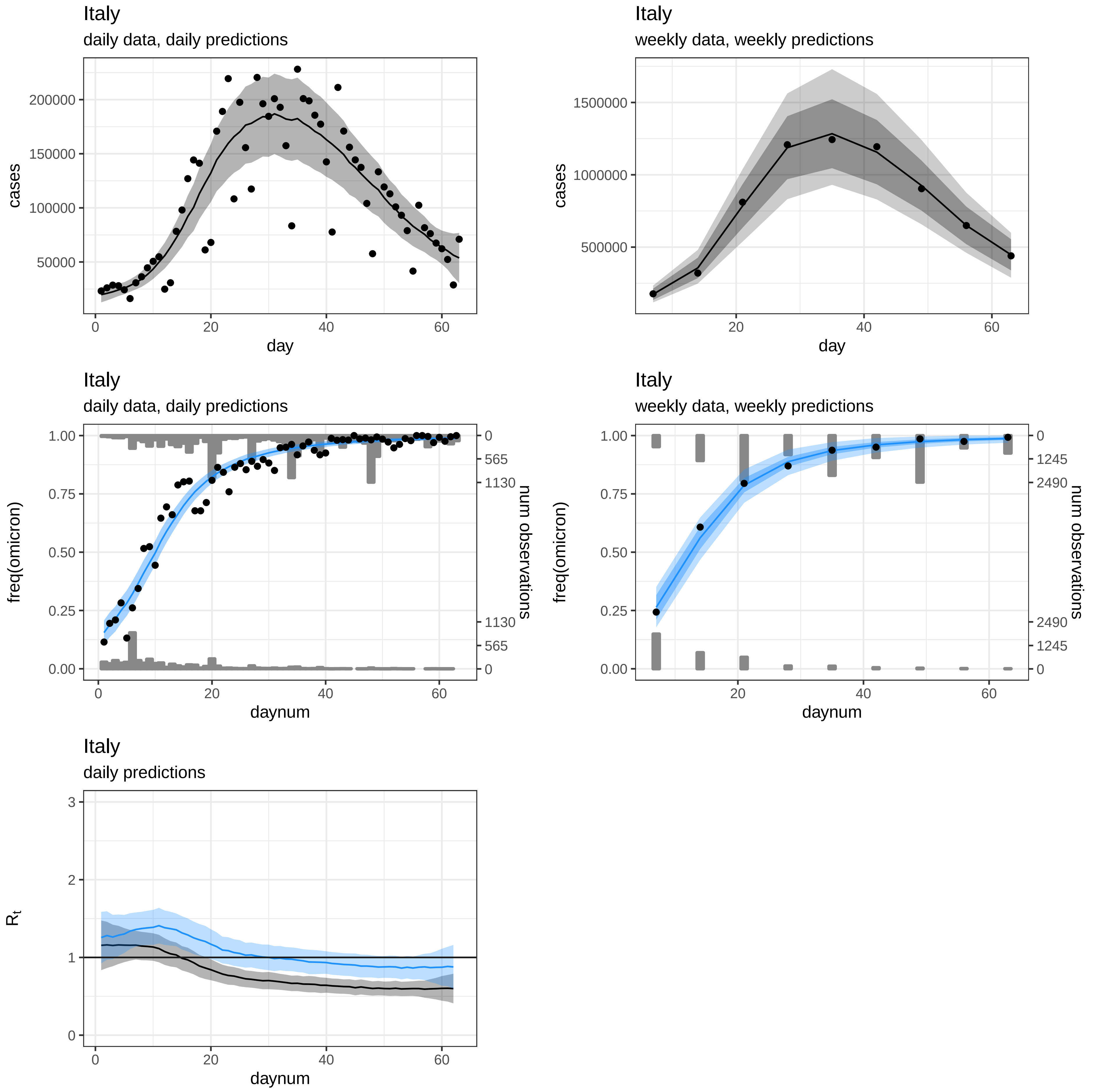
Italy
daily data, daily predictions
cases
day
200000
150000
100000
50000
0
20
40
60
Italy
weekly data, weekly predictions
cases
day
1500000
1000000
500000
20
40
60
Italy
daily data, daily predictions
freq(omicron)
num observations
daynum
1.00
0.75
0.50
0.25
0.00
0
565
1130
1130
565
0
0
20
40
60
Italy
weekly data, weekly predictions
freq(omicron)
num observations
daynum
1.00
0.75
0.50
0.25
0.00
0
1245
2490
2490
1245
0
20
40
60
Italy
daily predictions
$R_t$
daynum
3
2
1
0
0
20
40
60

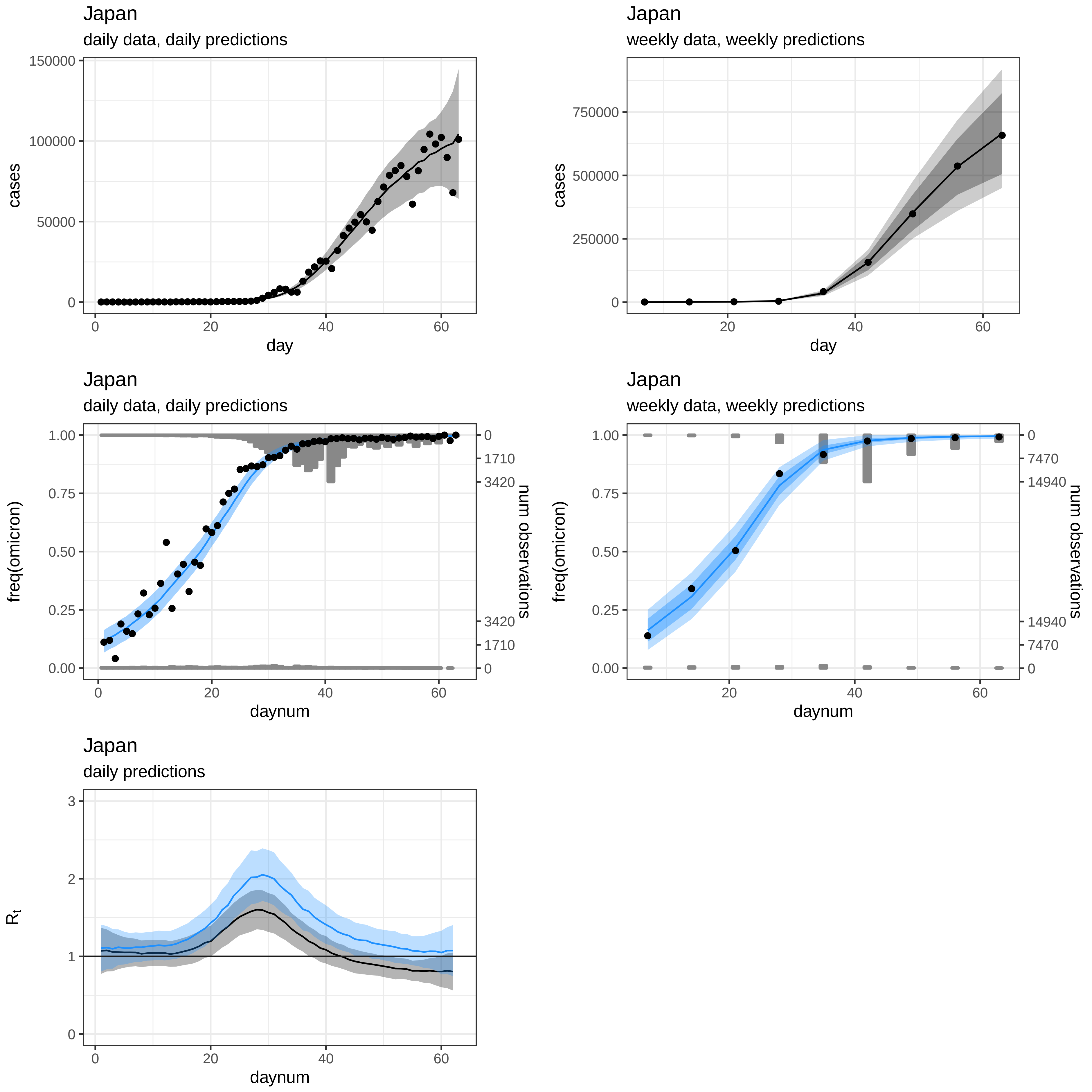
Japan
daily data, daily predictions
cases
150000
100000
50000
0
0
20
40
60
day
Japan
weekly data, weekly predictions
cases
750000
500000
250000
0
20
40
60
day
Japan
daily data, daily predictions
freq(omicron)
1.00
0.75
0.50
0.25
0.00
0
1710
3420
3420
1710
0
num observations
0
20
40
60
daynum
Japan
weekly data, weekly predictions
freq(omicron)
1.00
0.75
0.50
0.25
0.00
0
7470
14940
14940
7470
0
num observations
20
40
60
daynum
Japan
daily predictions
$R_t$
3
2
1
0
0
20
40
60
daynum

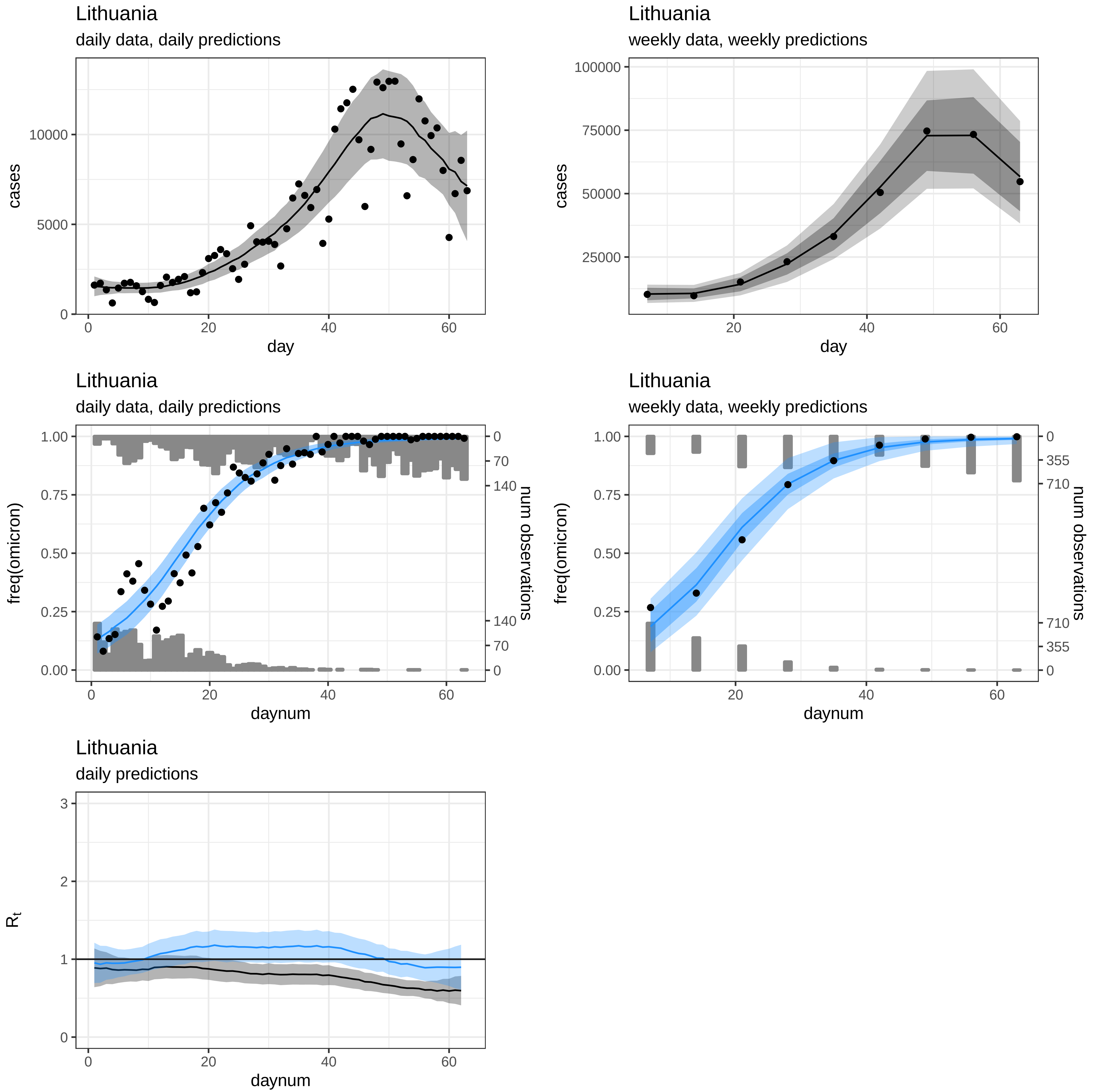
Lithuania
daily data, daily predictions
cases
day
Lithuania
weekly data, weekly predictions
cases
day
Lithuania
daily data, daily predictions
freq(omicron)
num observations
daynum
Lithuania
weekly data, weekly predictions
freq(omicron)
num observations
daynum
Lithuania
daily predictions
$R_t$
daynum

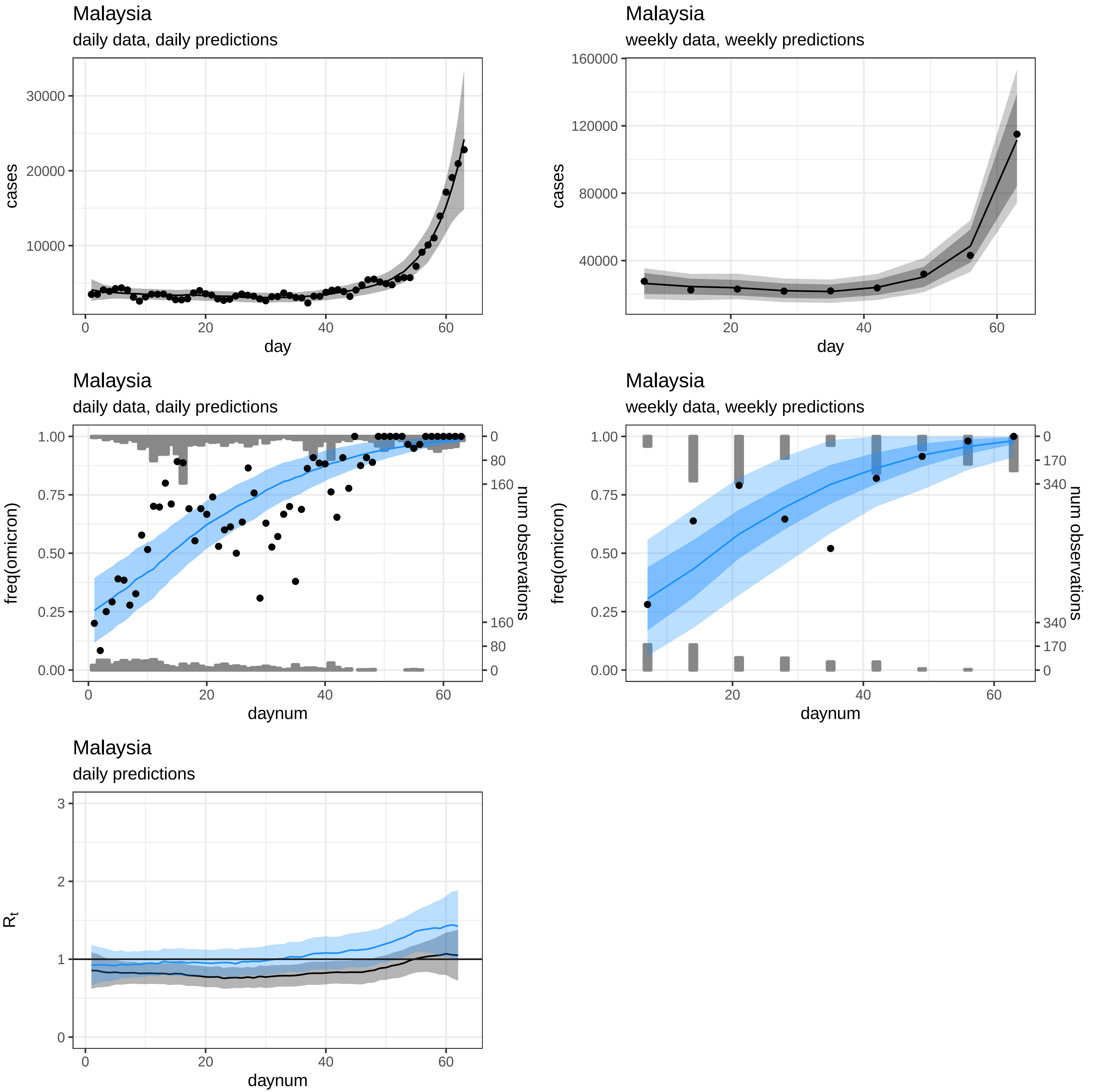
Malaysia
daily data, daily predictions
cases
day
Malaysia
weekly data, weekly predictions
cases
day
Malaysia
daily data, daily predictions
freq(omicron)
num observations
daynum
Malaysia
weekly data, weekly predictions
freq(omicron)
num observations
daynum
Malaysia
daily predictions
$R_t$
daynum

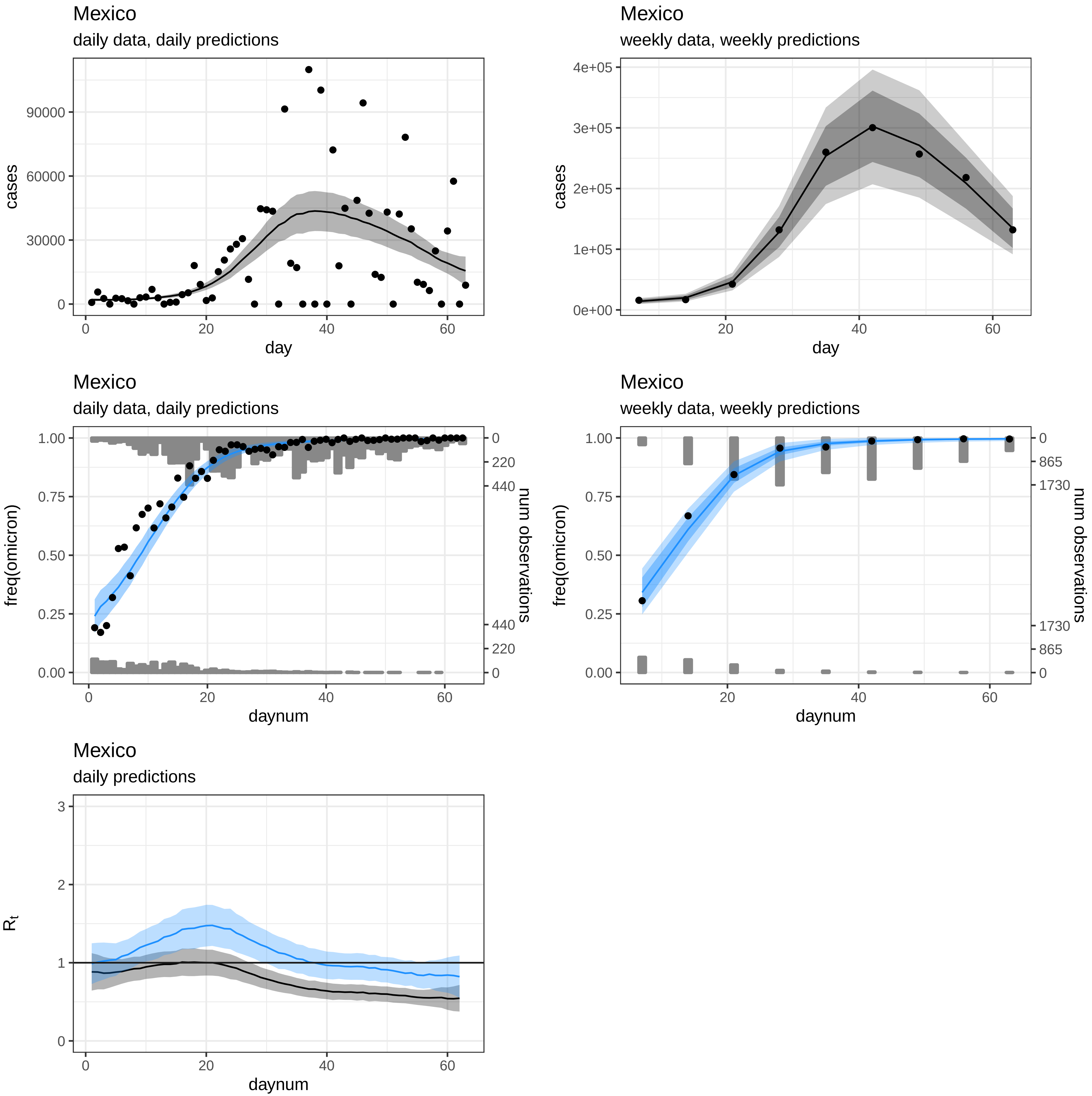
Mexico
daily data, daily predictions
cases
day
Mexico
weekly data, weekly predictions
cases
day
Mexico
daily data, daily predictions
freq(omicron)
num observations
daynum
Mexico
weekly data, weekly predictions
freq(omicron)
num observations
daynum
Mexico
daily predictions
$R_t$
daynum

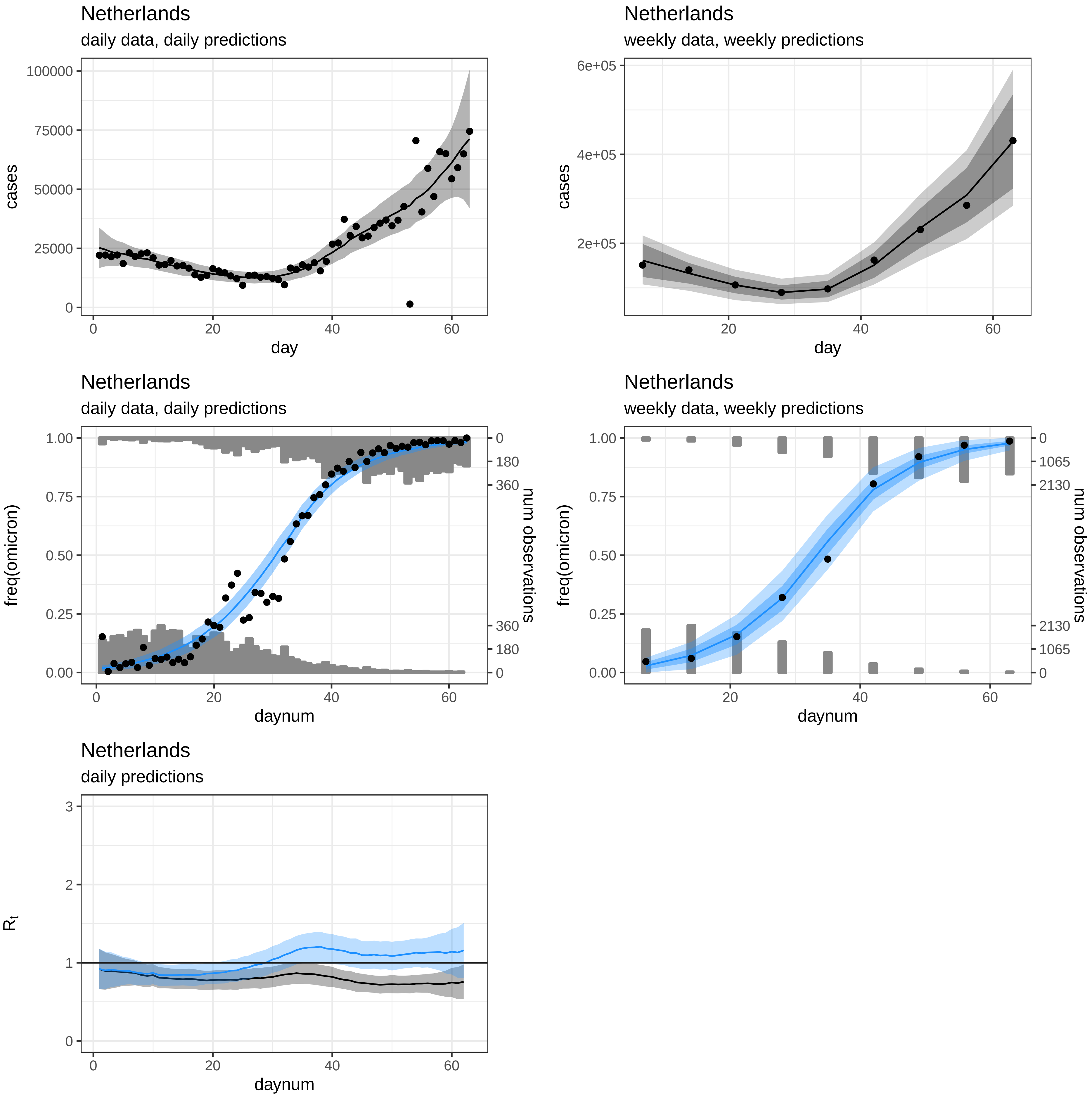
Netherlands
daily data, daily predictions
cases
day
Netherlands
weekly data, weekly predictions
cases
day
Netherlands
daily data, daily predictions
freq(omicron)
num observations
daynum
Netherlands
weekly data, weekly predictions
freq(omicron)
num observations
daynum
Netherlands
daily predictions
$R_t$
daynum

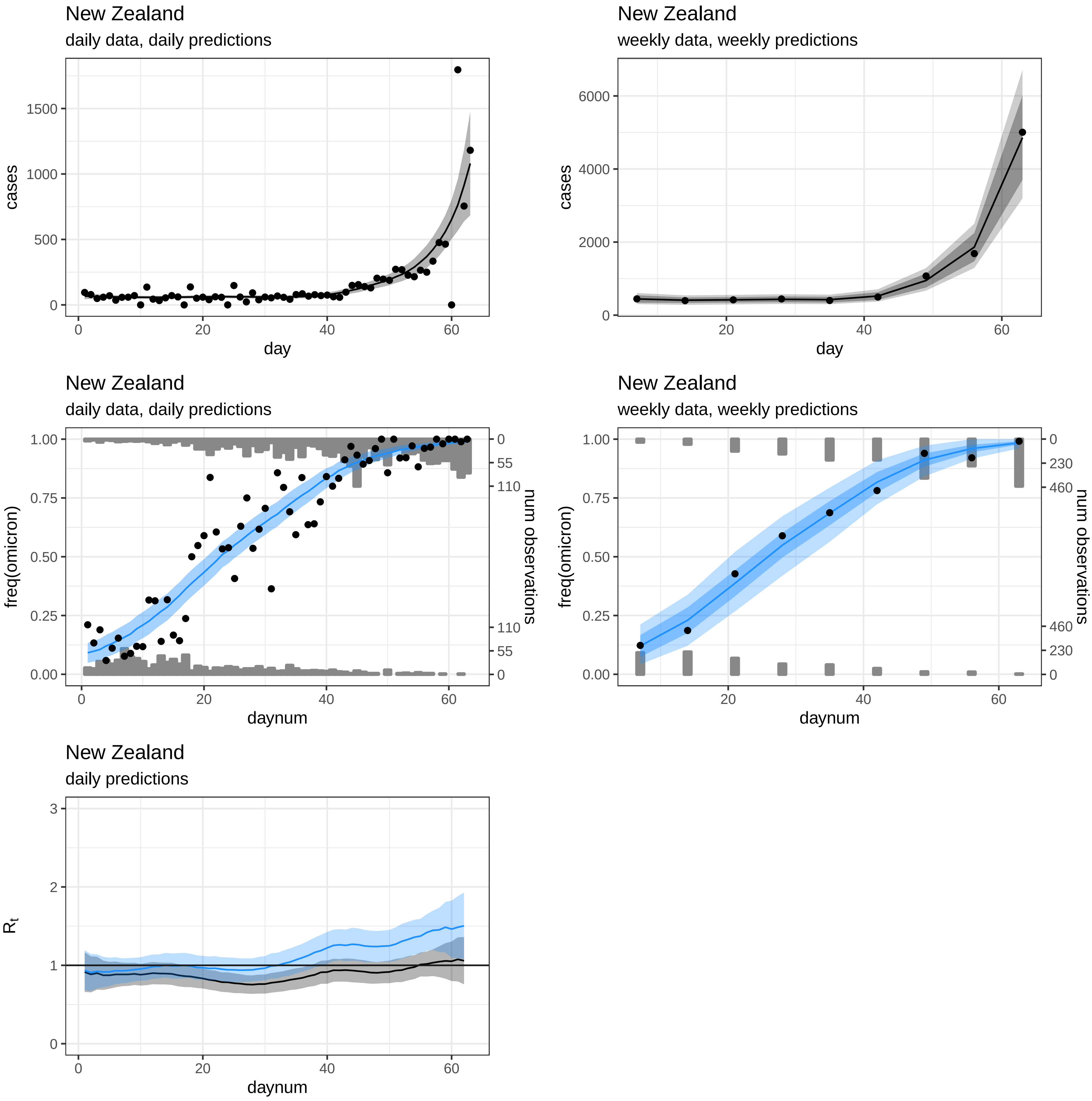
New Zealand
daily data, daily predictions
cases
day
New Zealand
weekly data, weekly predictions
cases
day
New Zealand
daily data, daily predictions
freq(omicron)
num observations
daynum
New Zealand
weekly data, weekly predictions
freq(omicron)
num observations
daynum
New Zealand
daily predictions
$R_t$
daynum

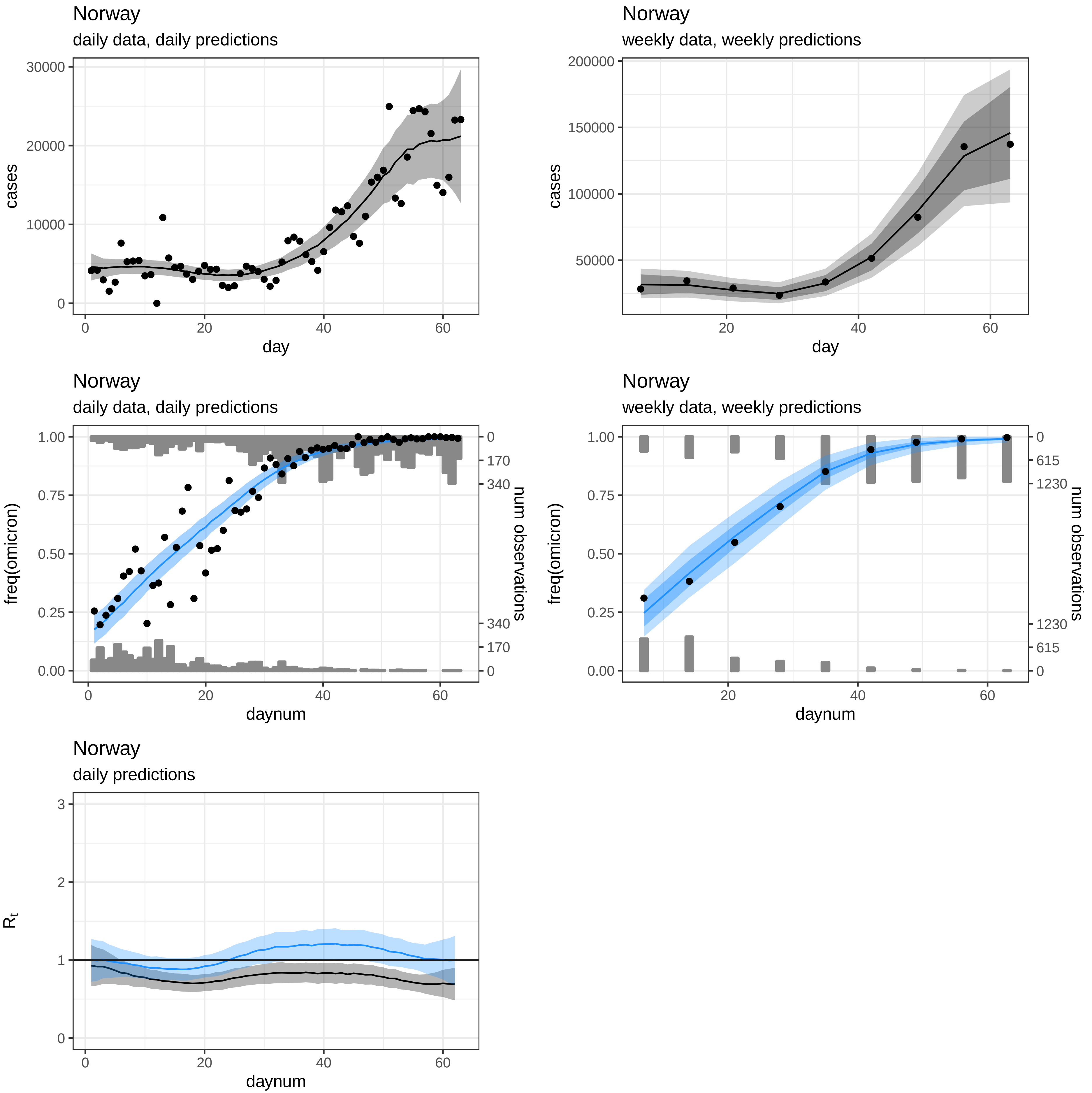
Norway
daily data, daily predictions
cases
day
Norway
weekly data, weekly predictions
cases
day
Norway
daily data, daily predictions
freq(omicron)
num observations
daynum
Norway
weekly data, weekly predictions
freq(omicron)
num observations
daynum
Norway
daily predictions
$R_t$
daynum

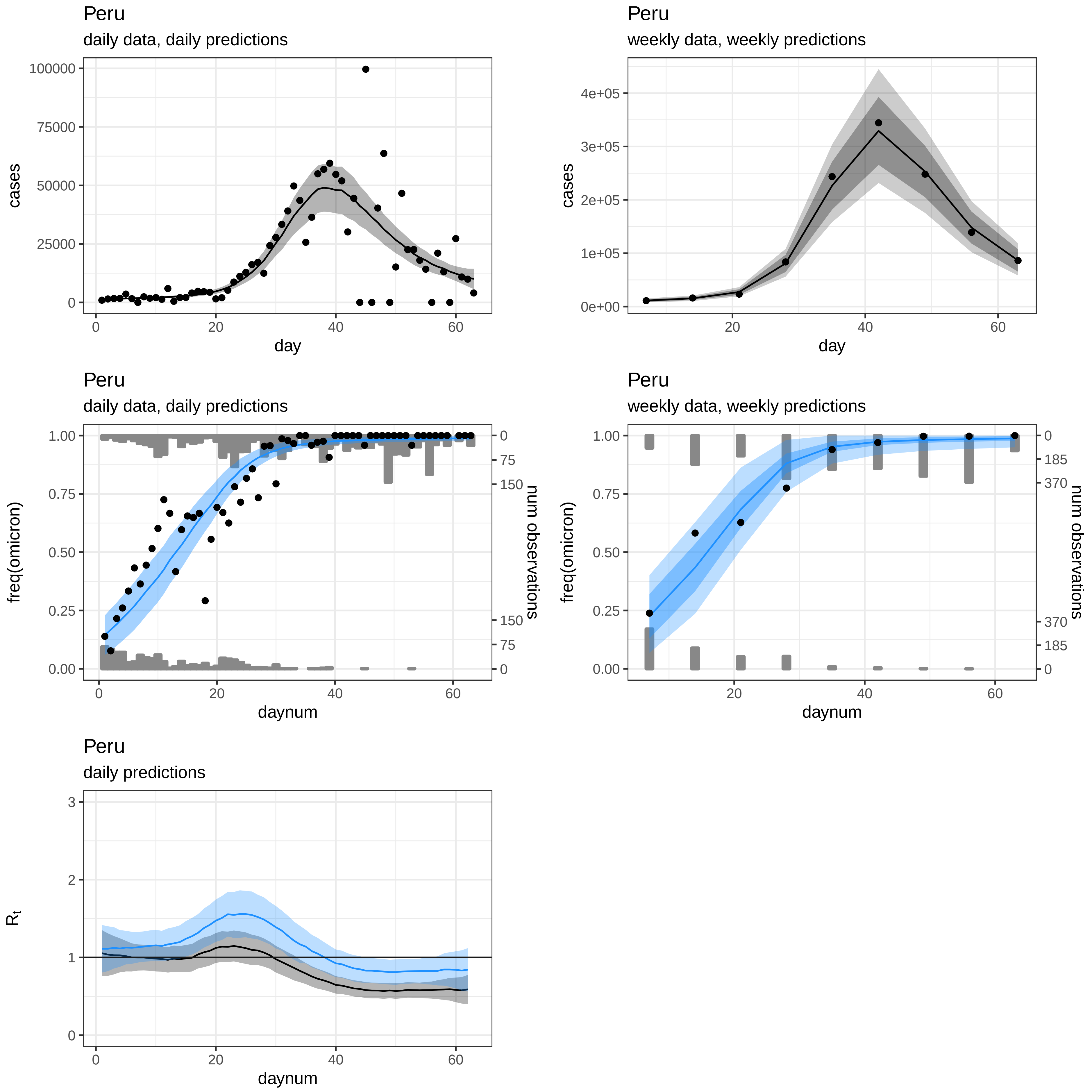

Peru
daily data, daily predictions
cases
100000
75000
50000
25000
0
0
20
40
60
day
Peru
weekly data, weekly predictions
cases
4e+05
3e+05
2e+05
1e+05
0e+00
20
40
60
day
Peru
daily data, daily predictions
freq(omicron)
1.00
0.75
0.50
0.25
0.00
0
75
150
150
75
0
num observations
0
20
40
60
daynum
Peru
weekly data, weekly predictions
freq(omicron)
1.00
0.75
0.50
0.25
0.00
0
185
370
370
185
0
num observations
20
40
60
daynum
Peru
daily predictions
$R_t$
3
2
1
0
0
20
40
60
daynum

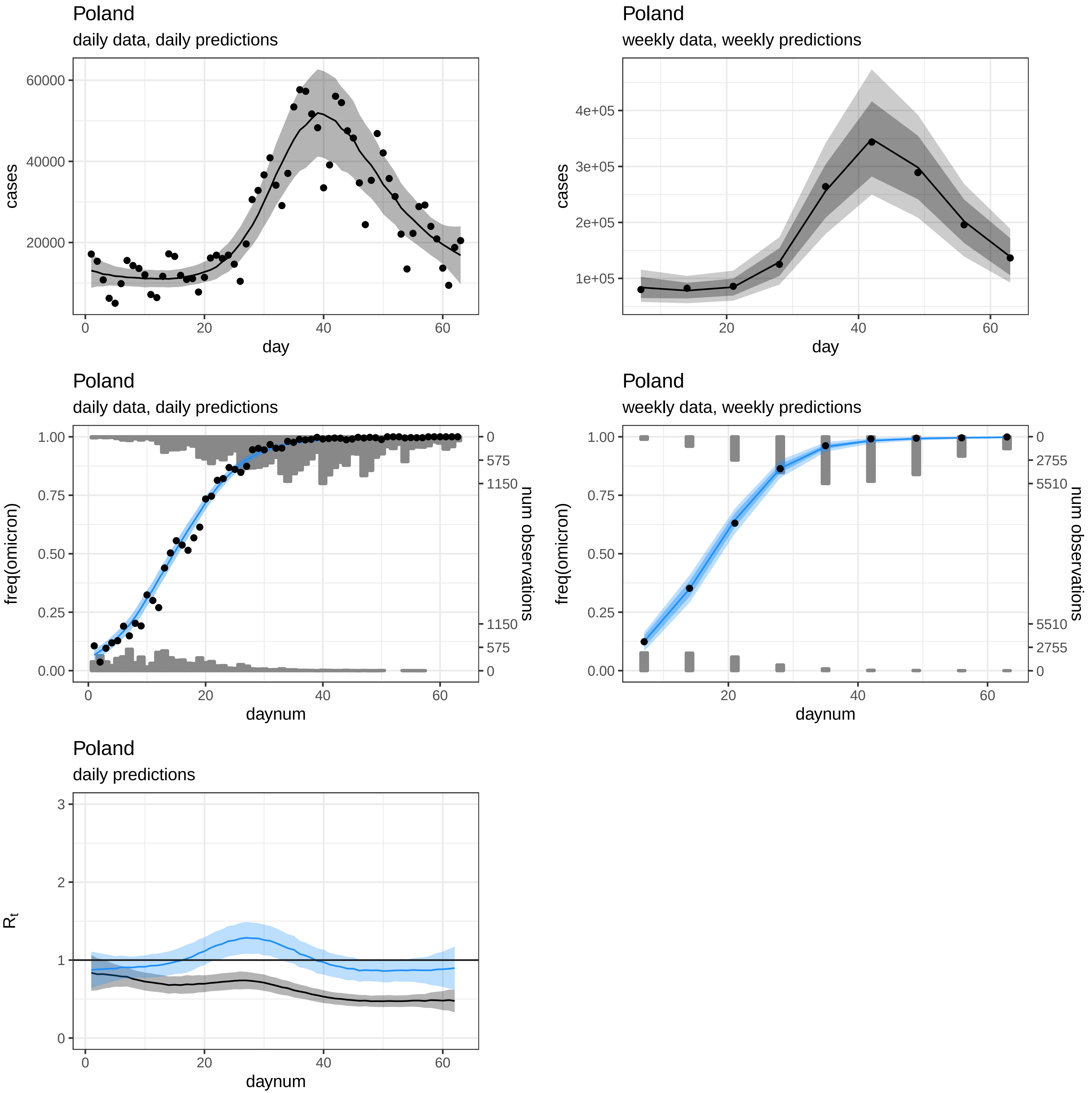

Poland
daily data, daily predictions
cases
day
Poland
weekly data, weekly predictions
cases
day
Poland
daily data, daily predictions
freq(omicron)
num observations
daynum
Poland
weekly data, weekly predictions
freq(omicron)
num observations
daynum
Poland
daily predictions
$R_t$
daynum

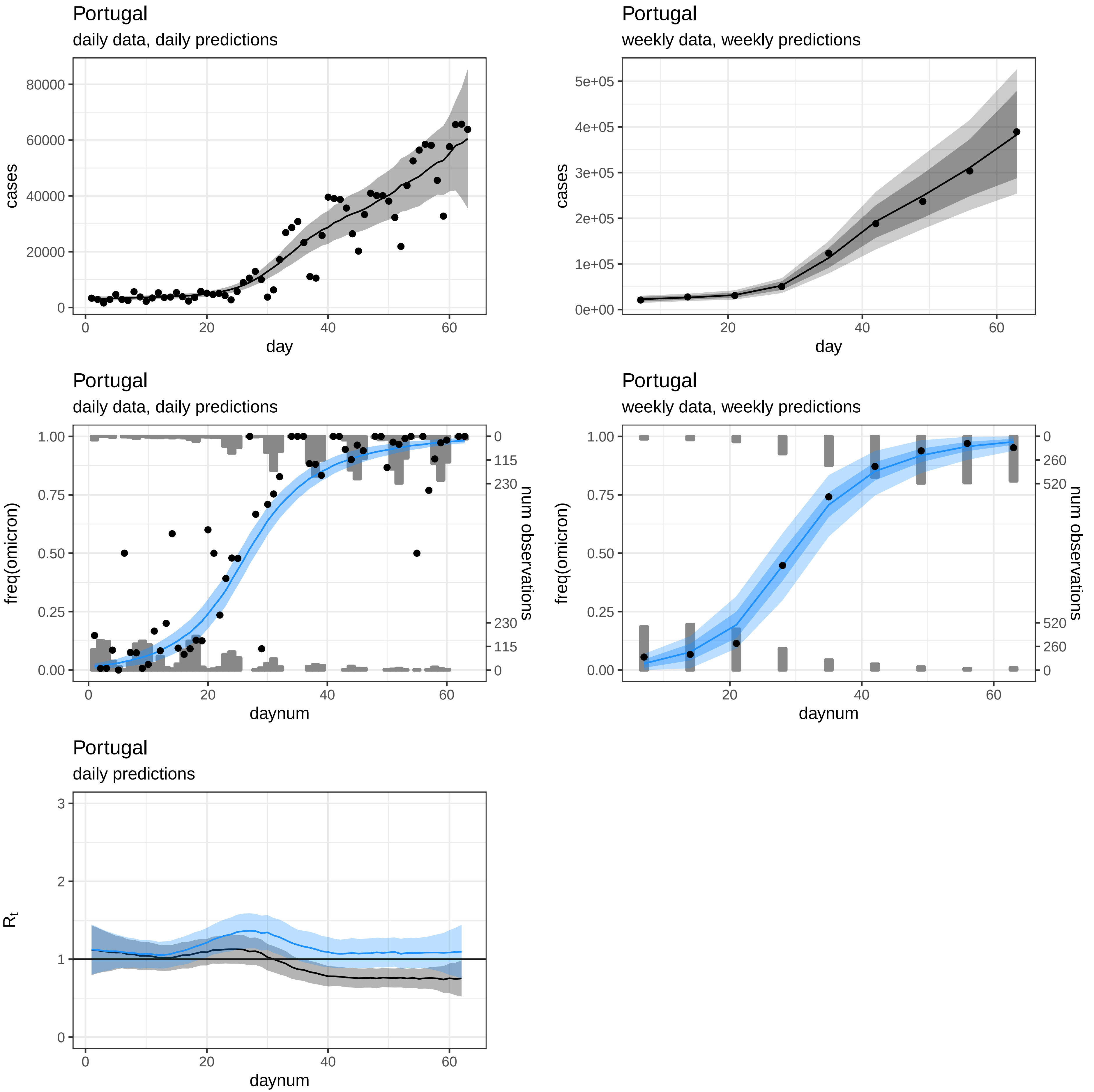
Portugal
daily data, daily predictions
cases
day
Portugal
weekly data, weekly predictions
cases
day
Portugal
daily data, daily predictions
freq(omicron)
num observations
daynum
Portugal
weekly data, weekly predictions
freq(omicron)
num observations
daynum
Portugal
daily predictions
$R_t$
daynum

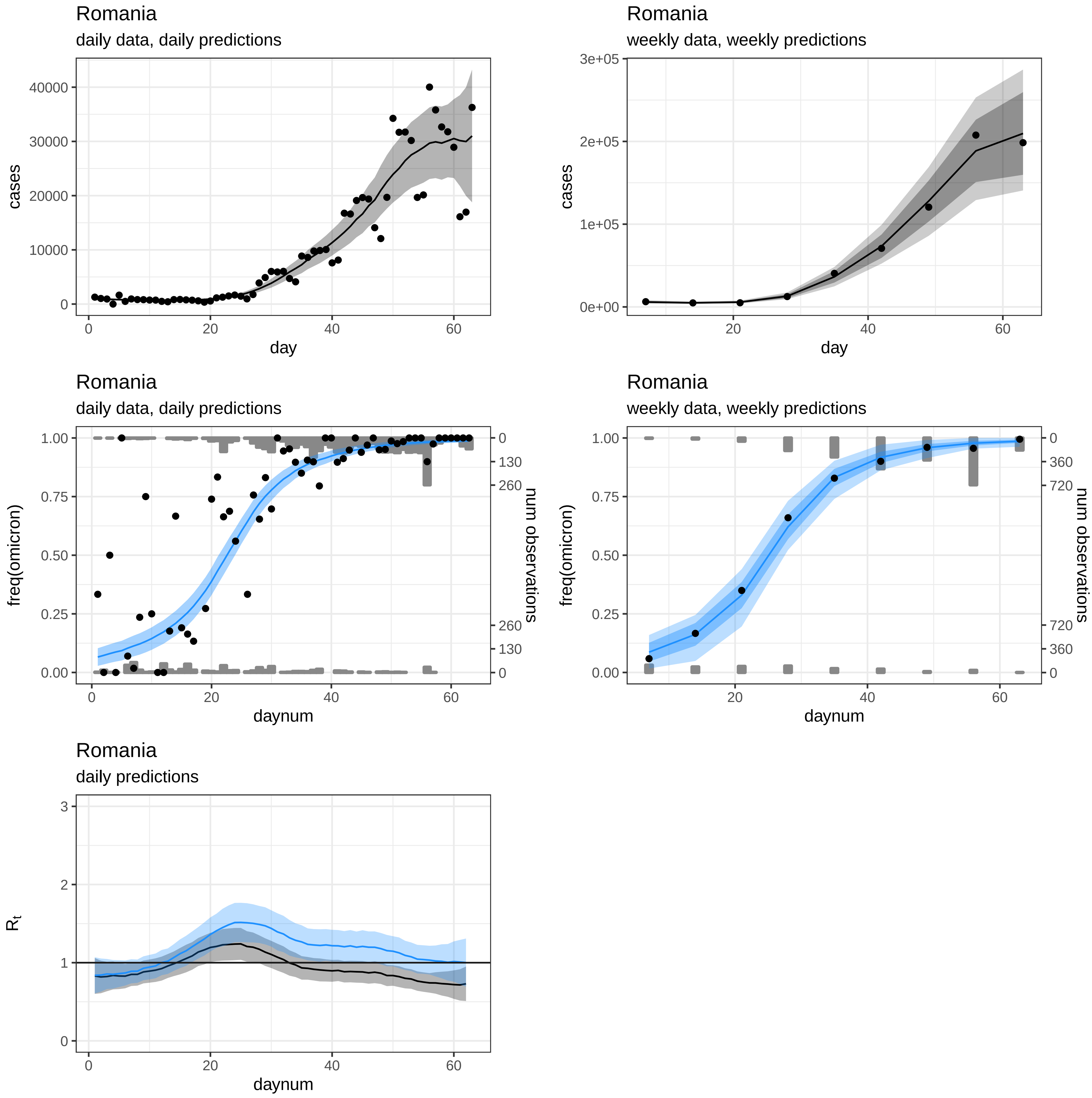
Romania
daily data, daily predictions
cases
day
Romania
weekly data, weekly predictions
cases
day
Romania
daily data, daily predictions
freq(omicron)
num observations
daynum
Romania
weekly data, weekly predictions
freq(omicron)
num observations
daynum
Romania
daily predictions
$R_t$
daynum

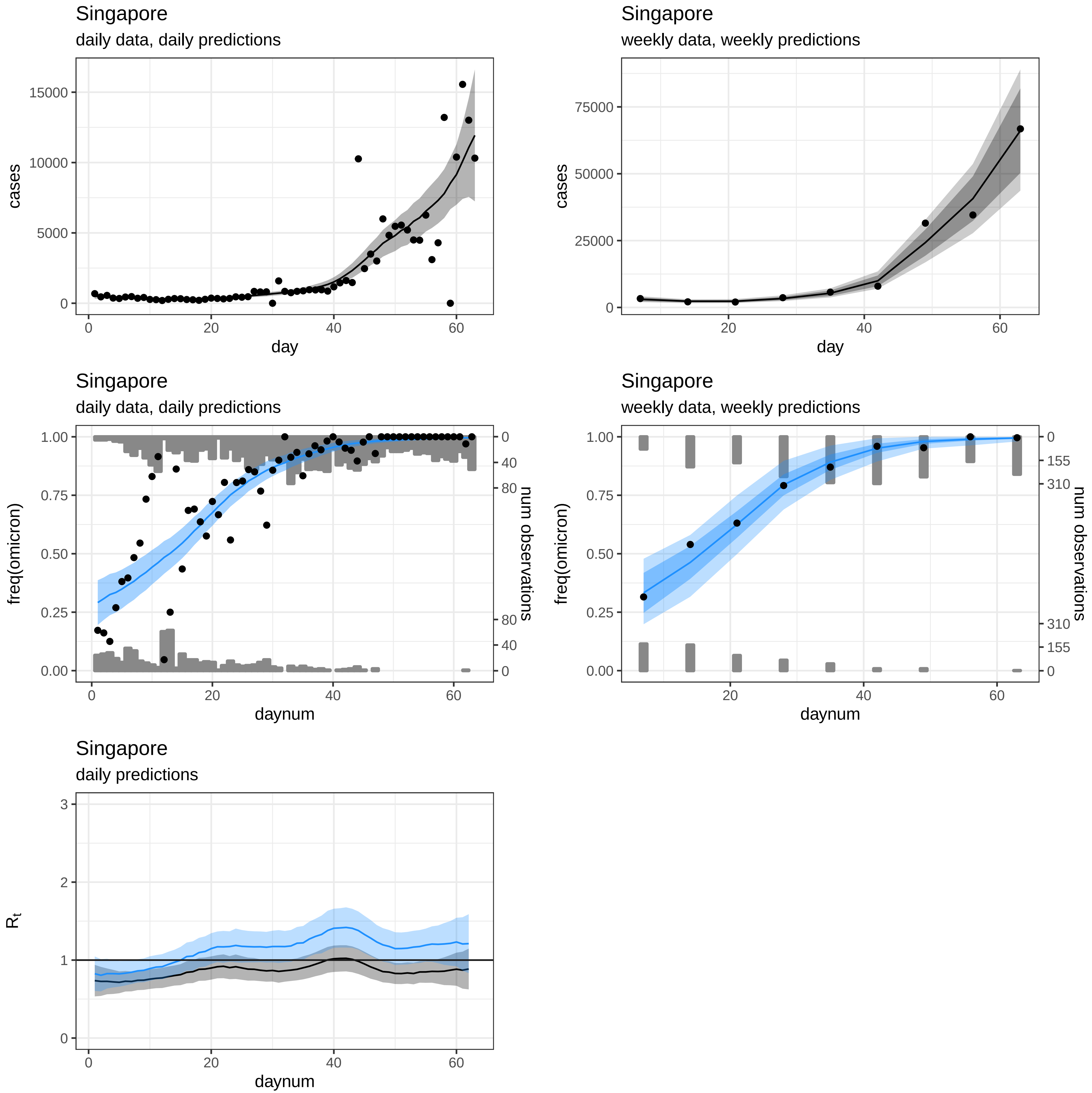

Singapore
daily data, daily predictions
cases
day
Singapore
weekly data, weekly predictions
cases
day
Singapore
daily data, daily predictions
freq(omicron)
num observations
daynum
Singapore
weekly data, weekly predictions
freq(omicron)
num observations
daynum
Singapore
daily predictions
$R_t$
daynum

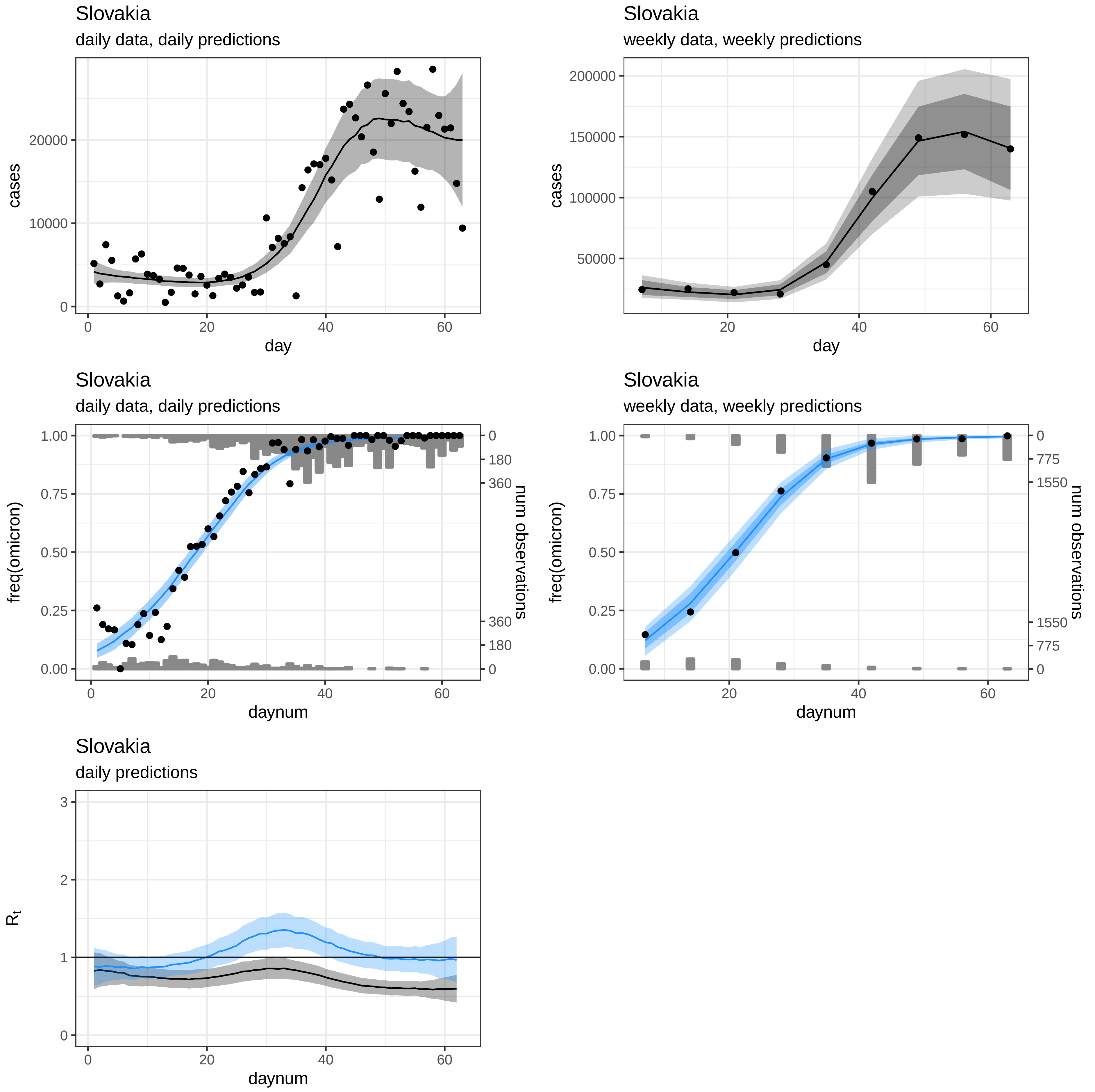

Slovakia
daily data, daily predictions
cases
0
10000
20000
day
0
20
40
60
Slovakia
weekly data, weekly predictions
cases
50000
100000
150000
200000
day
20
40
60
Slovakia
daily data, daily predictions
freq(omicron)
0.00
0.25
0.50
0.75
1.00
num observations
0
180
360
360
180
0
daynum
0
20
40
60
Slovakia
weekly data, weekly predictions
freq(omicron)
0.00
0.25
0.50
0.75
1.00
num observations
0
775
1550
1550
775
0
daynum
20
40
60
Slovakia
daily predictions
$R_t$
0
1
2
3
daynum
0
20
40
60

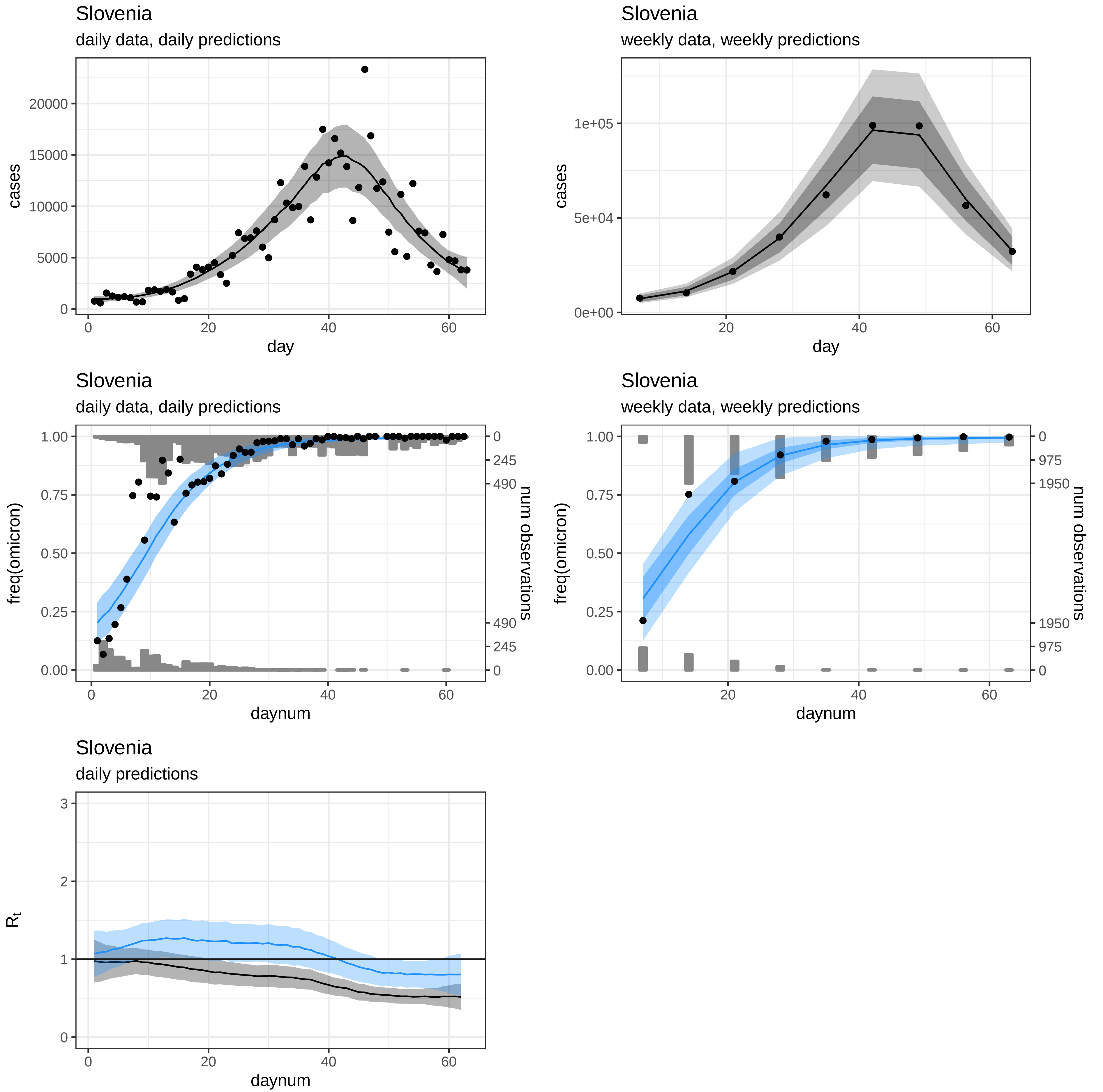
Slovenia
daily data, daily predictions
cases
day
Slovenia
weekly data, weekly predictions
cases
day
Slovenia
daily data, daily predictions
freq(omicron)
num observations
daynum
Slovenia
weekly data, weekly predictions
freq(omicron)
num observations
daynum
Slovenia
daily predictions
$R_t$
daynum

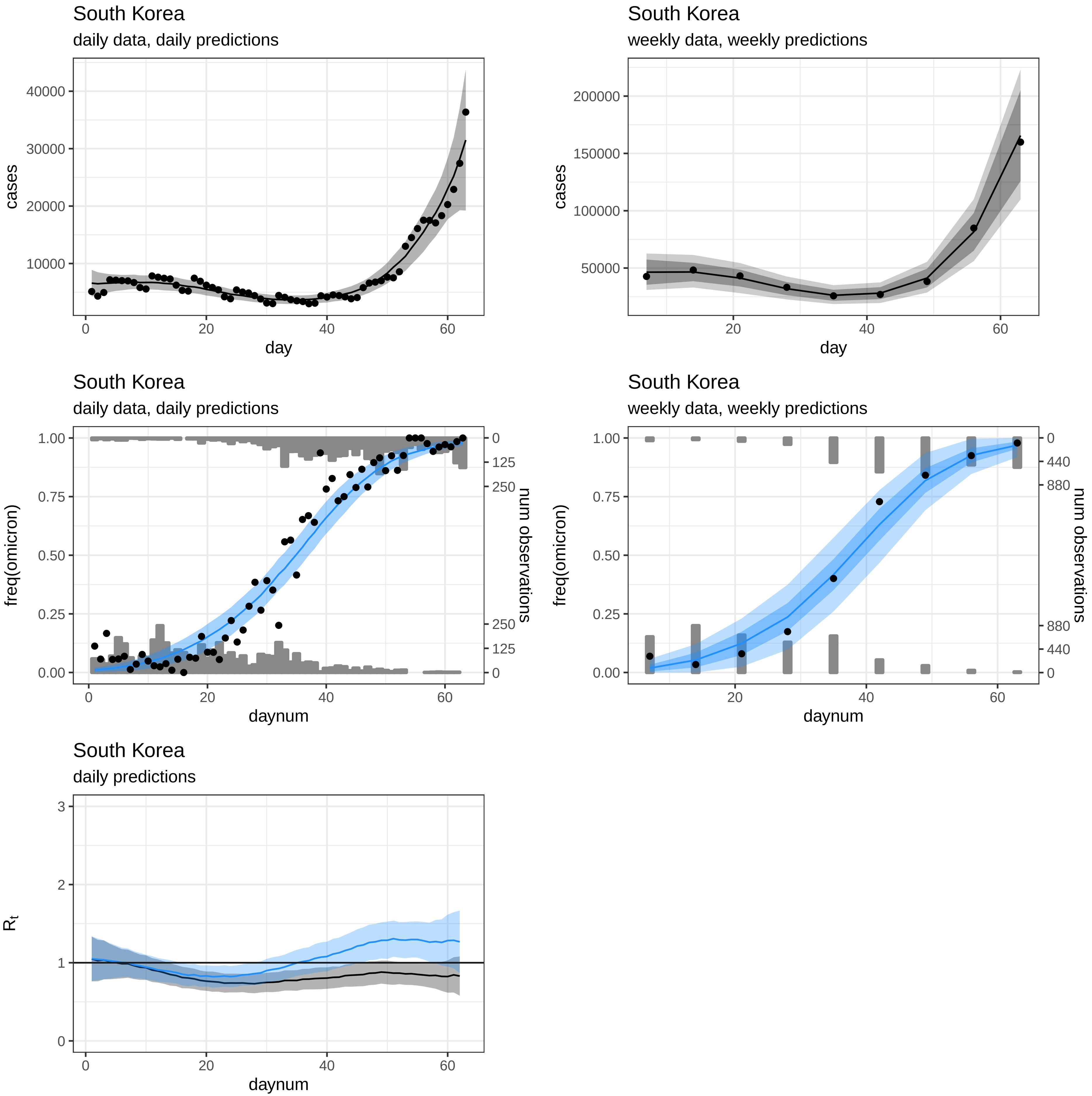
South Korea
daily data, daily predictions
cases
40000
30000
20000
10000
0
20
40
60
day
South Korea
weekly data, weekly predictions
cases
200000
150000
100000
50000
20
40
60
day
South Korea
daily data, daily predictions
freq(omicron)
1.00
0.75
0.50
0.25
0.00
num observations
0
125
250
250
125
0
0
20
40
60
daynum
South Korea
weekly data, weekly predictions
freq(omicron)
1.00
0.75
0.50
0.25
0.00
num observations
0
440
880
880
440
0
20
40
60
daynum
South Korea
daily predictions
$R_t$
3
2
1
0
0
20
40
60
daynum

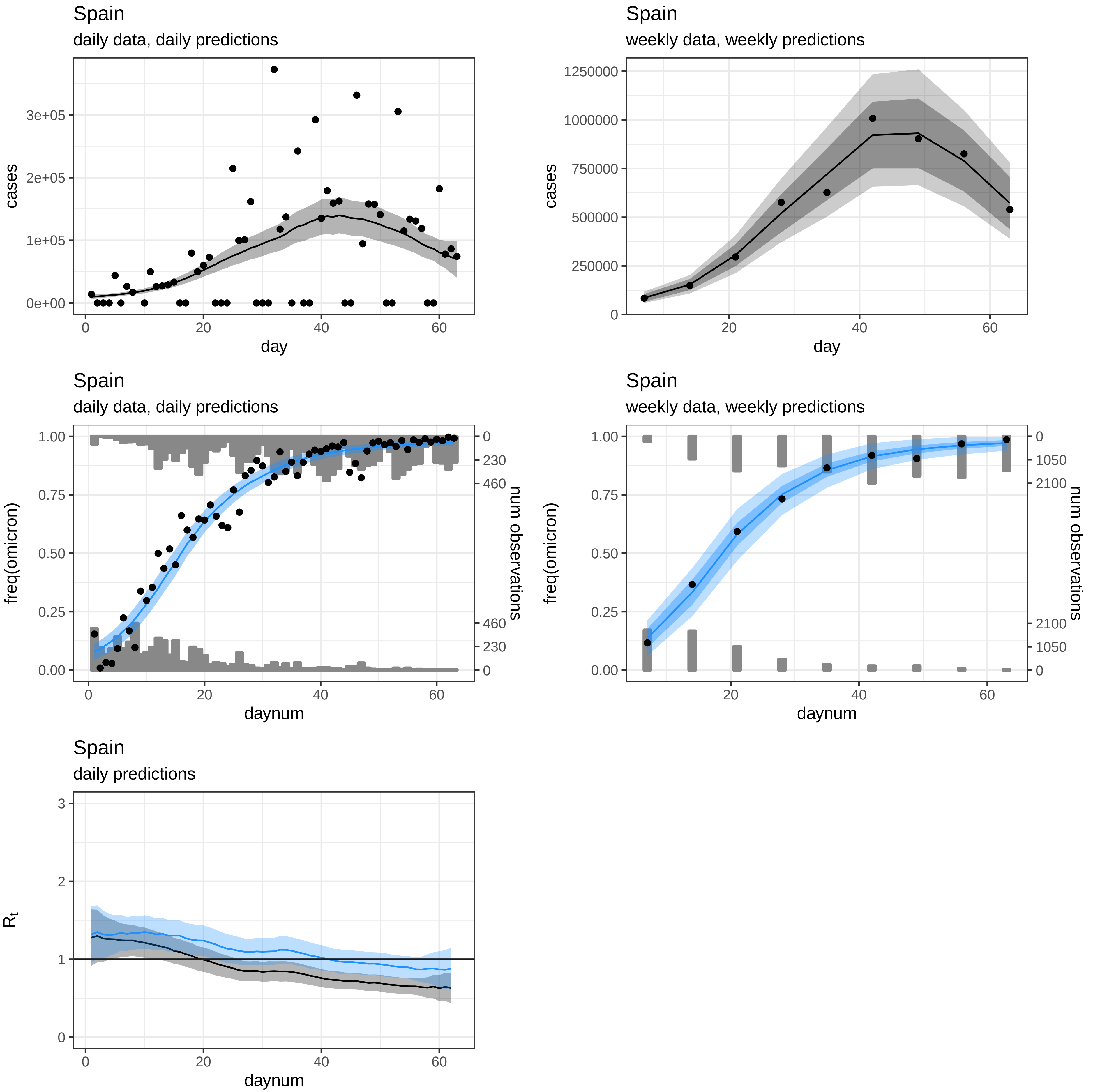
Spain
daily data, daily predictions
cases
3e+05
2e+05
1e+05
0e+00
0
20
40
60
day
Spain
weekly data, weekly predictions
cases
1250000
1000000
750000
500000
250000
0
20
40
60
day
Spain
daily data, daily predictions
freq(omicron)
1.00
0.75
0.50
0.25
0.00
num observations
0
230
460
460
230
0
0
20
40
60
daynum
Spain
weekly data, weekly predictions
freq(omicron)
1.00
0.75
0.50
0.25
0.00
num observations
0
1050
2100
2100
1050
0
20
40
60
daynum
Spain
daily predictions
$R_t$
3
2
1
0
0
20
40
60
daynum

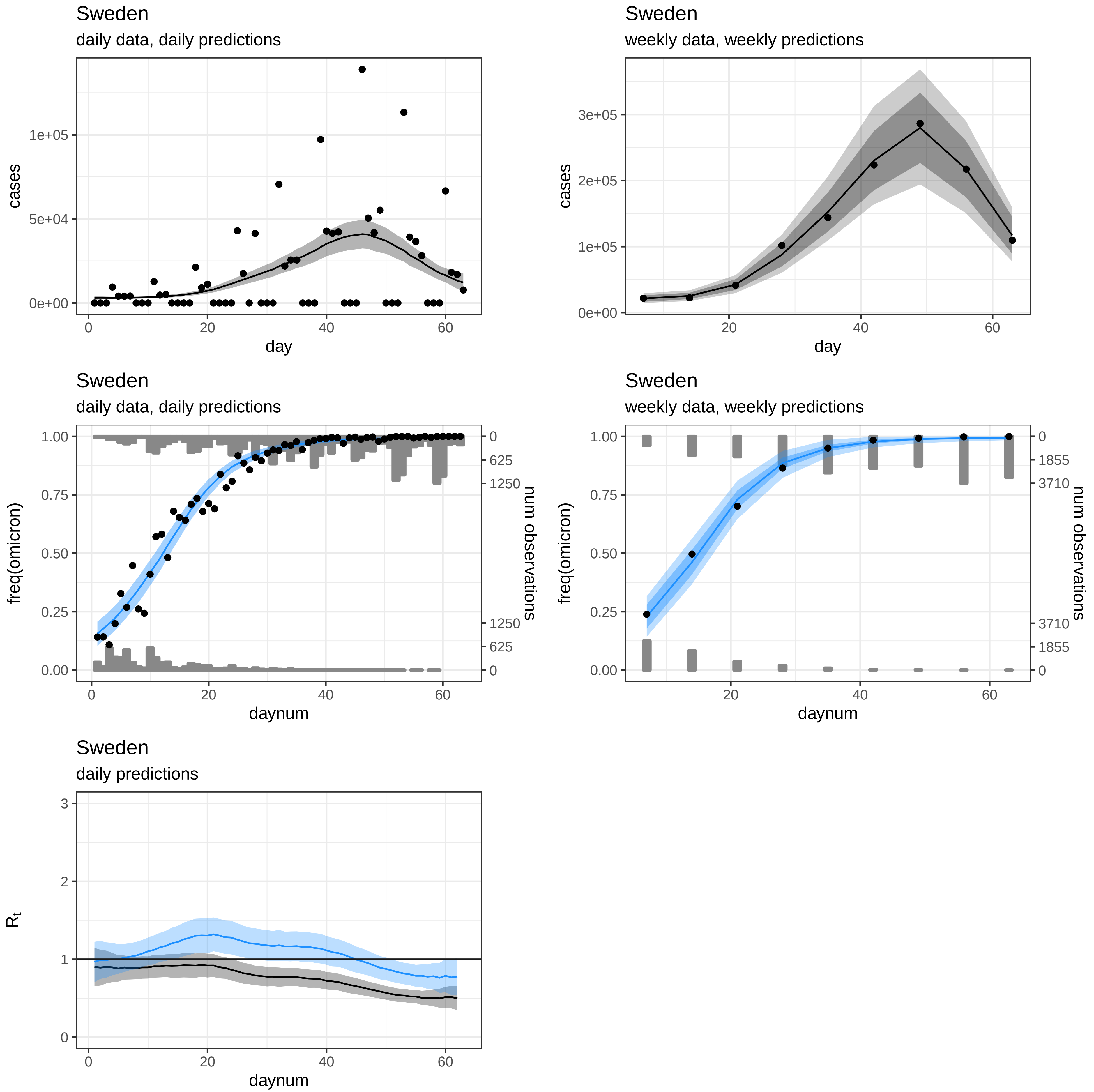
Sweden
daily data, daily predictions
cases
day
Sweden
weekly data, weekly predictions
cases
day
Sweden
daily data, daily predictions
freq(omicron)
num observations
daynum
Sweden
weekly data, weekly predictions
freq(omicron)
num observations
daynum
Sweden
daily predictions
$R_t$
daynum

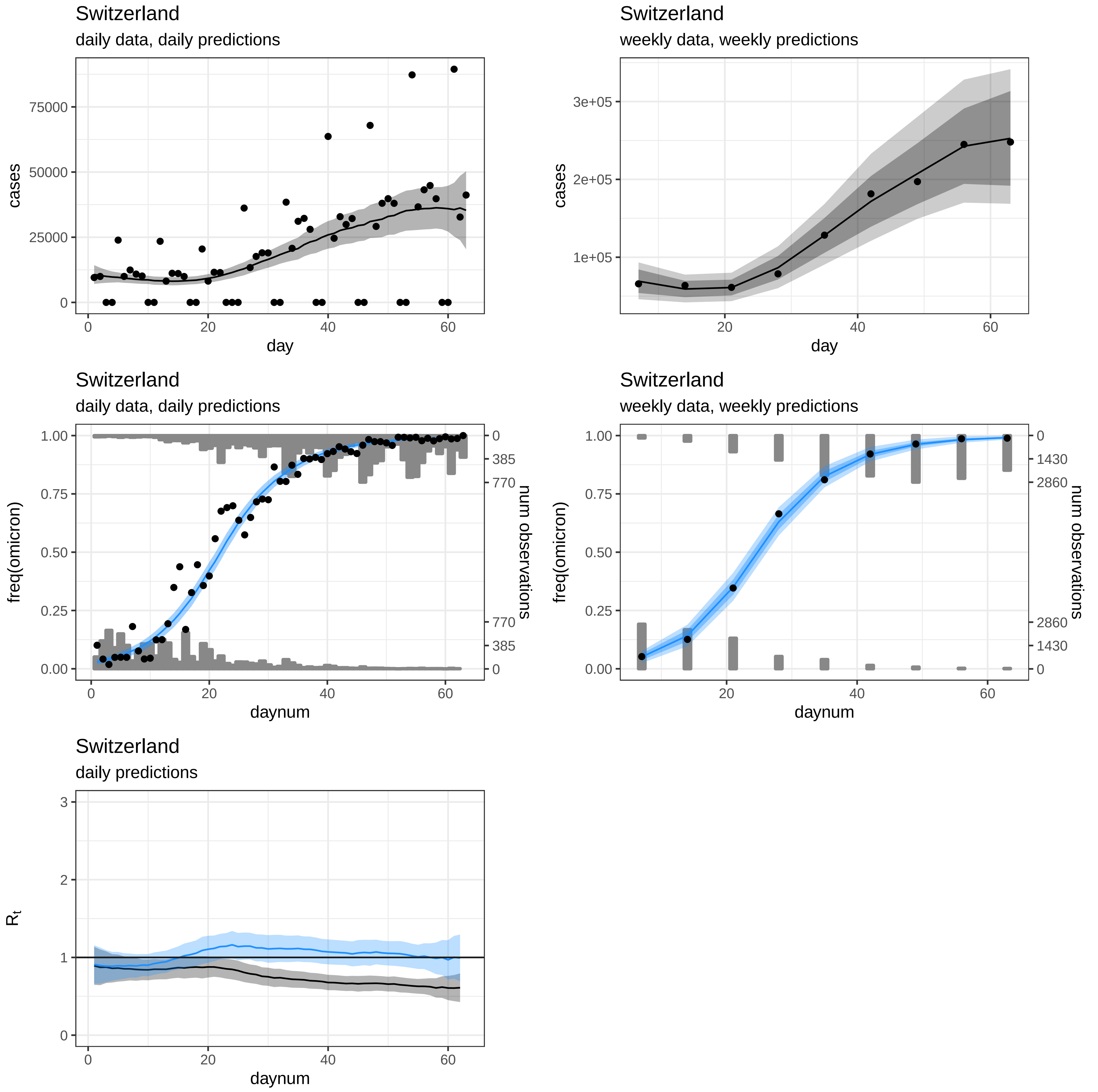

Switzerland
daily data, daily predictions
cases
day
Switzerland
weekly data, weekly predictions
cases
day
Switzerland
daily data, daily predictions
freq(omicron)
num observations
daynum
Switzerland
weekly data, weekly predictions
freq(omicron)
num observations
daynum
Switzerland
daily predictions
$R_t$
daynum

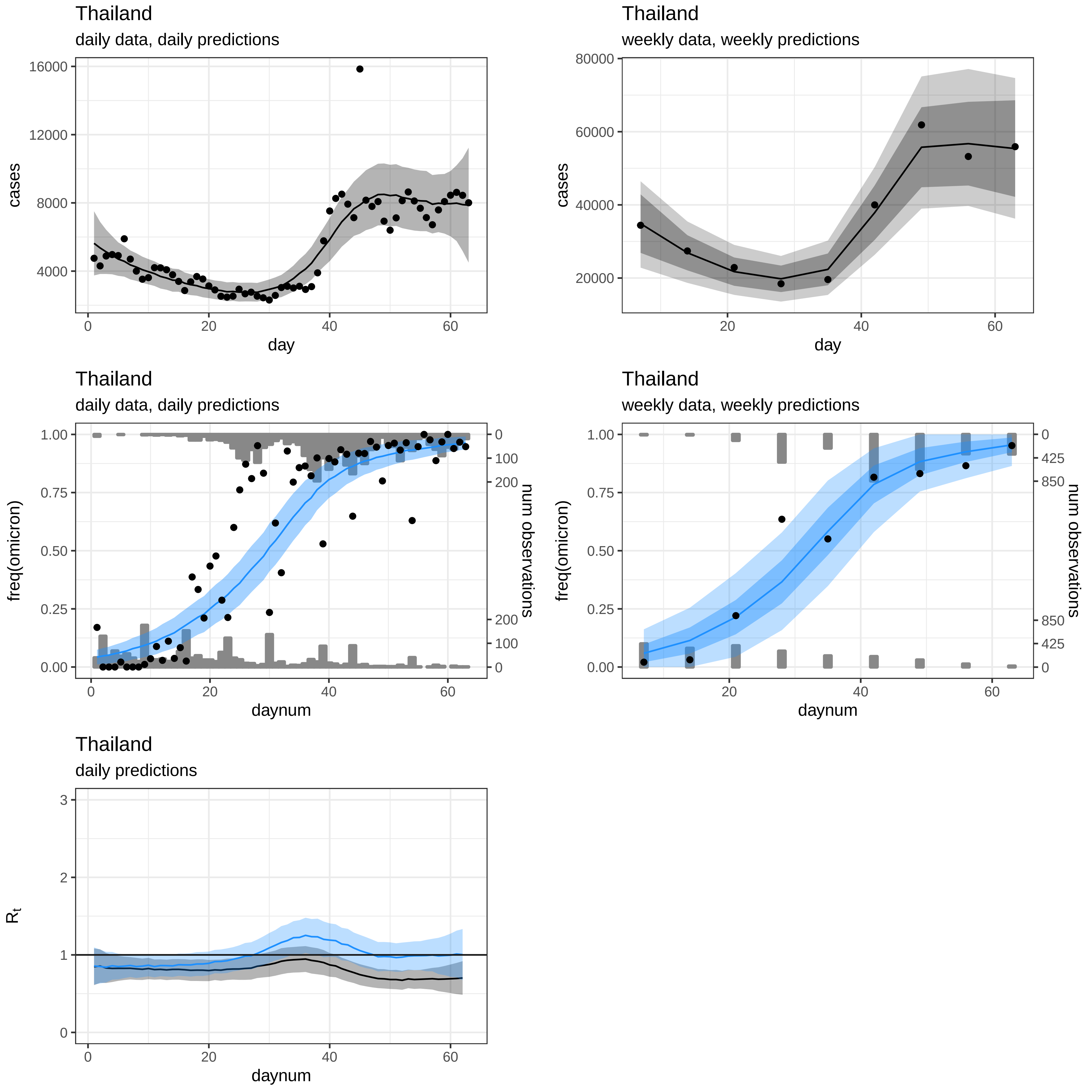
Thailand
daily data, daily predictions
cases
day
Thailand
weekly data, weekly predictions
cases
day
Thailand
daily data, daily predictions
freq(omicron)
num observations
daynum
Thailand
weekly data, weekly predictions
freq(omicron)
num observations
daynum
Thailand
daily predictions
$R_t$
daynum

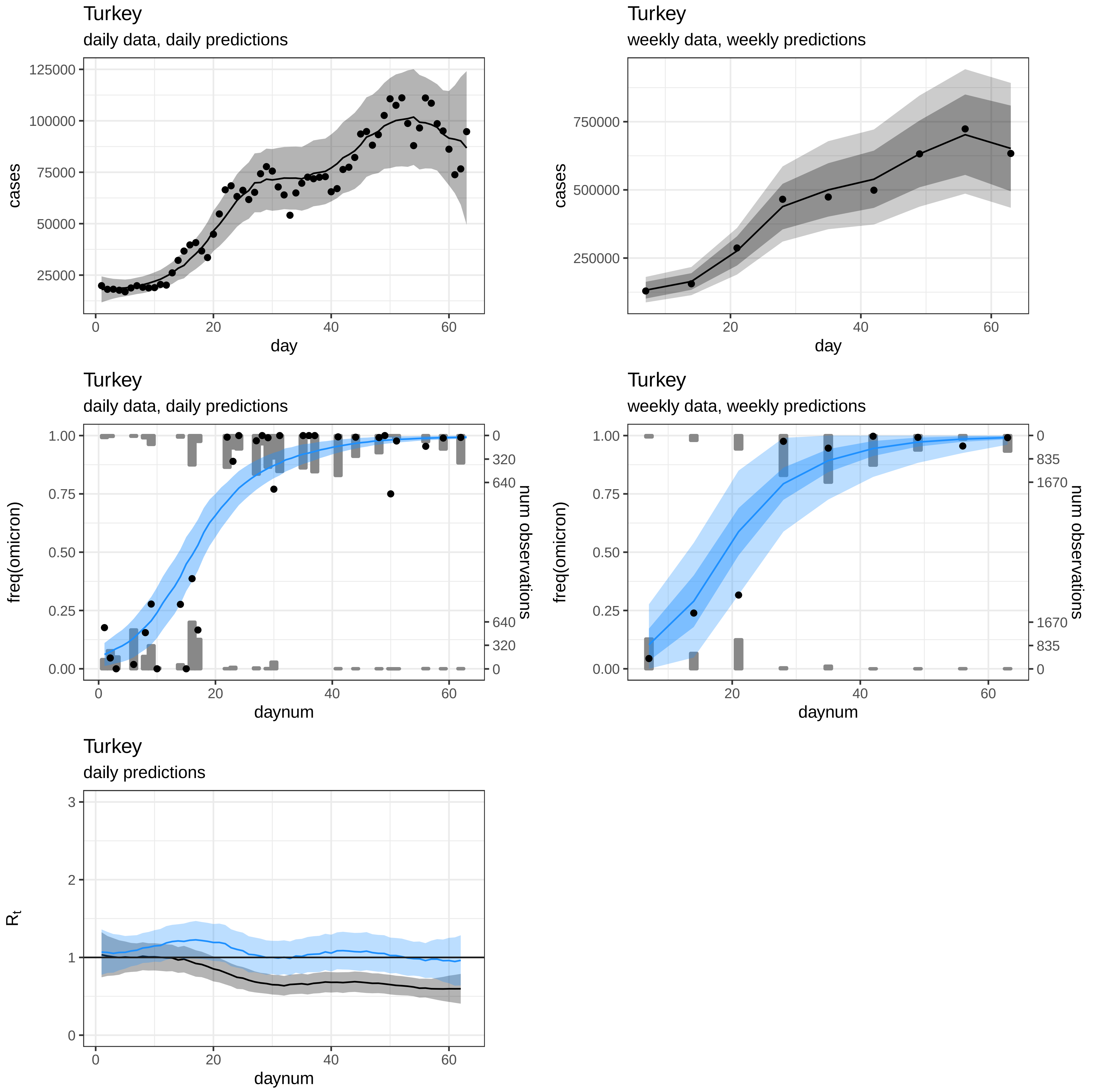
Turkey
daily data, daily predictions
cases
day
Turkey
weekly data, weekly predictions
cases
day
Turkey
daily data, daily predictions
freq(omicron)
num observations
daynum
Turkey
weekly data, weekly predictions
freq(omicron)
num observations
daynum
Turkey
daily predictions
$R_t$
daynum

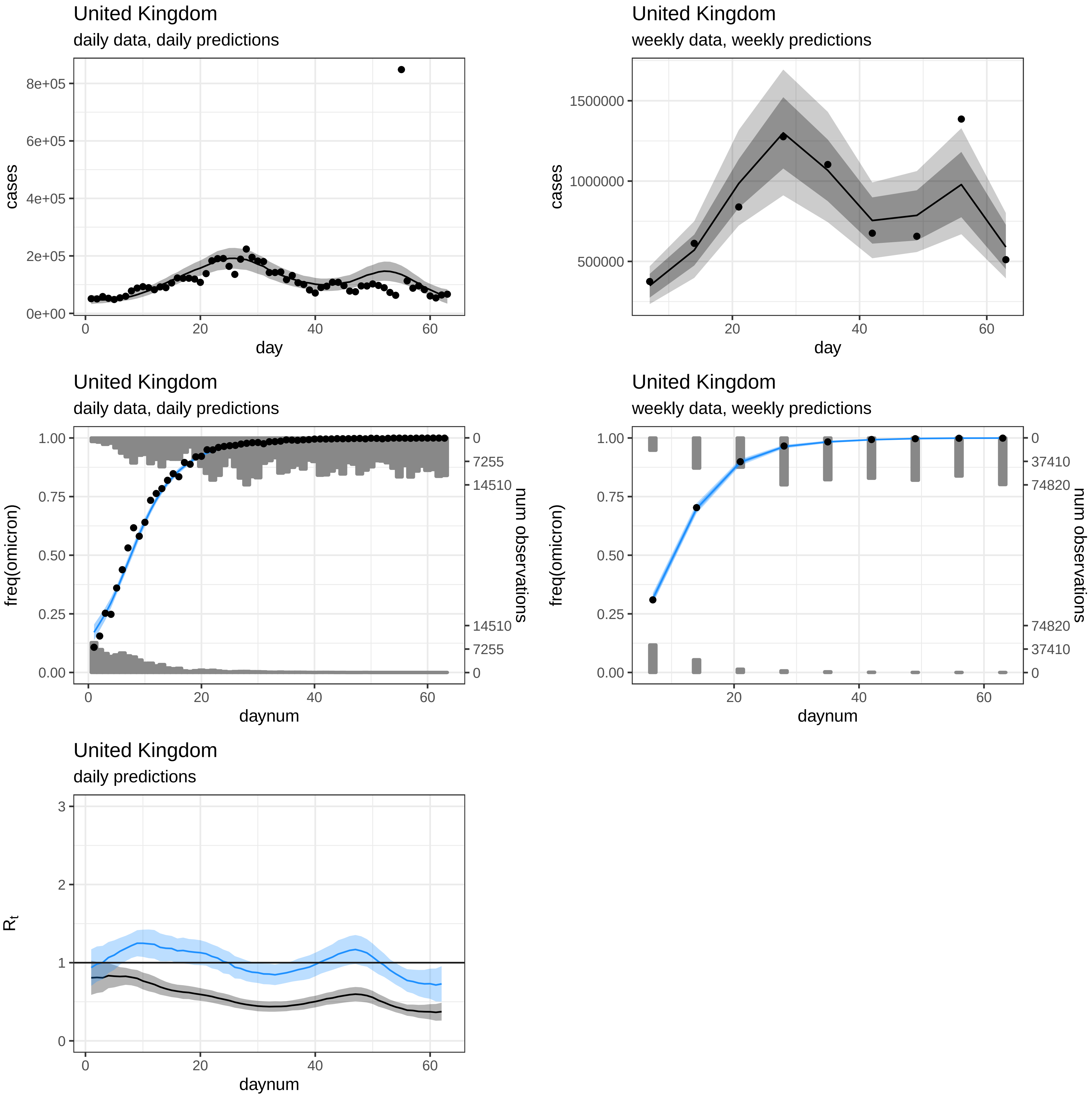
United Kingdom
daily data, daily predictions
8e+05
6e+05
4e+05
2e+05
0e+00
cases
0
20
40
60
day
United Kingdom
weekly data, weekly predictions
1500000
1000000
500000
cases
20
40
60
day
United Kingdom
daily data, daily predictions
1.00
0.75
0.50
0.25
0.00
freq(omicron)
0
7255
14510
14510
7255
0
num observations
0
20
40
60
daynum
United Kingdom
weekly data, weekly predictions
1.00
0.75
0.50
0.25
0.00
freq(omicron)
0
37410
74820
74820
37410
0
num observations
20
40
60
daynum
United Kingdom
daily predictions
3
2
1
0
$R_t$
0
20
40
60
daynum

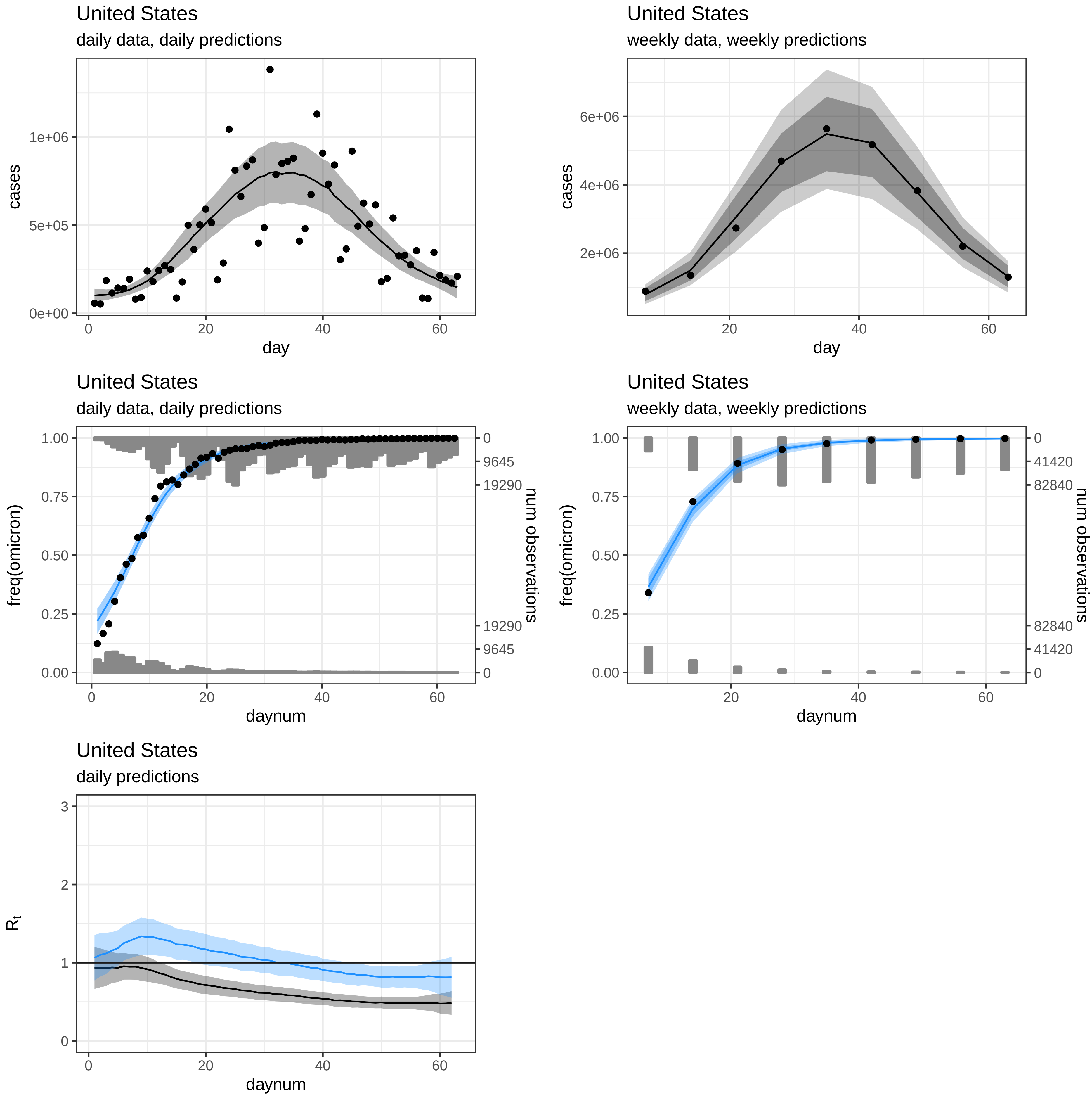
United States
daily data, daily predictions
cases
1e+06
5e+05
0e+00
0
20
40
60
day
United States
weekly data, weekly predictions
cases
6e+06
4e+06
2e+06
20
40
60
day
United States
daily data, daily predictions
freq(omicron)
1.00
0.75
0.50
0.25
0.00
0
9645
19290
num observations
0
20
40
60
daynum
United States
weekly data, weekly predictions
freq(omicron)
1.00
0.75
0.50
0.25
0.00
0
41420
82840
num observations
20
40
60
daynum
United States
daily predictions
$R_t$
3
2
1
0
0
20
40
60
daynum
